# Current polygenic risk scores are unlikely to exacerbate unfairness in cardiovascular disease risk prediction

**DOI:** 10.1101/2025.09.18.25336069

**Authors:** Claire Coffey, Scott C. Ritchie, Lisa Pennells, Angela M. Wood, Michael Inouye, Samuel A. Lambert

## Abstract

1

**Background:** Current cardiovascular disease (CVD) risk prediction models place many individuals in an intermediate risk category where clinical decision-making remains uncertain, highlighting a critical gap in precision prevention. Polygenic risk scores (PRS) represent a promising solution to enhance risk stratification in intermediate-risk groups by identifying individuals with high genetic risk; however, observed differences in performance across ancestry groups may cause health disparities. The emerging field of algorithmic fairness offers a principled framework to assess equity in model performance among relevant subgroups, but have rarely been applied to clinical risk tools and PRS.

**Objectives:** To evaluate the fairness of incorporating PRS in CVD risk prediction, both as a standalone risk factor and as a risk-enhancing factor for individuals at intermediate risk (recommended in current clinical consensus statements).

**Methods:** Using data from the UK Biobank (N = 327,923), we calculated 10-year CVD risk using QRISK3 (a guideline-endorsed prediction model) and quantified genetic risk using a validated PRS. We assessed fairness among population characteristics relevant for health equity (age, sex, ethnicity, and area-level deprivation) using four algorithmic fairness metrics relevant for prevention (accuracy equality, equal opportunity, conditional use accuracy equality, and treatment equality).

**Results:** PRS, when used as a stand-alone risk factor, demonstrated fairness levels similar to or better than traditional clinical predictors (age, sex, blood pressure, cholesterol). Some variation in fairness was observed across ethnic groups, especially at extreme risk thresholds. When integrated as a risk-enhancing factor for reclassifying intermediate-risk individuals into high-risk categories, PRS improved sensitivity of CVD risk prediction with minimal impact on fairness metrics across demographic groups.

**Conclusions:** This study demonstrates that PRS, when incorporated into existing risk prediction frameworks, are unlikely to meaningfully exacerbate disparities in CVD risk stratification. Applying algorithmic fairness metrics provides insight into the equitable implementation of PRS and supports current recommendations for their use in risk-stratification for intermediate-risk individuals.

## 2 Introduction

Cardiovascular disease (CVD) is the leading cause of preventable morbidity and mortality worldwide, as approximately 70% of CVD events are attributable to modifiable risk factors.[1] Primary prevention of CVD focuses on identifying high-risk individuals, often using conventional risk factors (age, sex, blood pressure, and cholesterol measurements) to predict 10-year CVD risk in order to stratify the population and identify the high-risk population most likely to benefit from treatment (therapeutic or lifestyle interventions).[2, 3] A remaining challenge is that a significant proportion of individuals undergoing risk assessments will be estimated to have intermediate risk, subsequent treatment decisions are based on shared decision making with clinicians informed by existing or novel risk factors. As such, there is great interest in identifying novel risk factors to improve risk-stratification.

Genetics explains a significant amount of population variation in who develops CVD (heritability = 50%),[4] and can be quantified using polygenic risk scores (PRSs). PRSs estimate an individual’s predisposition to a disease based on the cumulative effect of many genetic variants identified by genome-wide association studies (GWAS).[5–7] For CVD, PRSs have shown promise in improving risk stratification when combined with conventional risk factors.[8–11] Integration of PRSs may improve early detection and prevention, particularly among younger individuals with high genetic risk who may not be captured by traditional models.[12–16] PRSs are increasingly being trialled in clinical settings, including in the UK and US.[17–21] Although not standard of care, clinical consensus statements have identified that PRSs are a risk factor for CVD with potential utility, particularly when applied as a risk-enhancing factor to reclassify intermediate risk individuals to a higher risk category that is likely to benefit from treatment.[22, 23]

The goal of risk stratification is to target treatment to improve outcomes, and should avoid creating disparities. Some clinical prediction models, including those for predicting CVD risk, have come under scrutiny for unfairness across population groups, often underperforming in individuals from ethnic minority or socio-economically disadvantaged backgrounds.[24–26] These disparities may arise from unrepresentative training data, systemic biases embedded in electronic health records, or failure to evaluate model performance across diverse groups.[27] Despite their promise, concerns have also been raised regarding the equity of PRS predictions.[28] Many PRS have a reduced prediction ability in individuals of non-European ancestry, a phenomenon known as the “transferability gap”. This is due to the limited ancestral diversity in GWAS datasets used to derive PRSs, which results in poorer predictive performance and potential disparities in clinical outcomes among ancestry groups.[29]

Algorithmic fairness is an expanding research area within the machine learning field, focusing on the development and application of mathematical definitions to quantify and promote fair predictions from models. It is notably used in high-stakes applications of machine learning (ML) [30], biostatistics, and artificial intelligence (AI), especially in domains including medical technology and risk prediction.[31–33] While mathematical definitions of fairness cannot independently eliminate societal discrimination and inequality, their measurement can identify biases across relevant demographic groups facilitating potential mitigations. Group fairness is typically assessed for key demographics in the population of interest that are often protected characteristics enshrined in law, including age, sex and ethnicity.[34, 35] Often other evaluated characteristics include proxies of social determinants of health; one of such example is area-level deprivation, which is also associated with the incidence of CVD.[36] Using fairness metrics to evaluate prediction results can be informative, illustrating cases where models have not accounted for group differences in risk, or groups that may require different thresholds for treatment decisions.

As PRSs move toward clinical use, it is critical to evaluate whether their integration exacerbates existing disparities. In this study, we apply metrics from the emerging field of algorithmic fairness to assess the fairness of using PRSs for CVD risk stratification at two levels. First, we evaluate the use of PRSs as a stand-alone risk factor for identifying high-risk individuals, contextualising their fairness in comparison to other commonly used CVD risk factors. Second, we measure the fairness of a guideline-recommended risk tool (QRISK3[3]) and assess the implications of applying PRSs to intermediate risk individuals in a two-stage stratification approach.

## 3 Methods

### 3.1 Study and participant data

This study used the UK Biobank (UKB), a longitudinal cohort of over 500,000 individuals aged 40–69 recruited between 2006 and 2010 via the UK National Health Service [37]. Participants underwent extensive baseline assessments and long-term follow-up, including questionnaires, nurses interview, biometrics, blood biomarkers, genotyping, and linkage to health records.

#### Fairness sub-groups

We selected age, sex, ethnicity, and area-level deprivation as the characteristics of interest for our fairness analysis based on their availability in UKB data, known associations with CVD, or status as protected characteristics that are of interest for addressing health inequalities and impacting health policy [36, 38, 39]. Age, sex and ethnicity data were extracted from the self-reported questionnaire data, and area-level deprivation was quantified by country-specific Index of Multiple Deprivation (IMD) from linked-participant data. Within each characteristic we defined sub-groups necessary to evaluate group fairness metrics: age, (40-*<*50, 50-*<*60 and 60-*<*70 years); ethnicity (self-reported White, South Asian, Black, and Other ethnic groups); sex; and quintiles of IMD scores (within the whole of UKB).

#### Clinical risk factors

Additional clinical risk factors required to calculate the QRISK3 CVD prediction algorithm were also extracted.[3] Physical measurements (body mass index [BMI], systolic blood pressure [SBP]) were taken from baseline assessment data, as well low-density, high-density, and total cholesterol measurements were extracted from the blood analysis. Medication use (BP-lowering, lipid-lowering, atypical antipsychotic) and lifestyle measurements (smoking, family history of CVD) were extracted from questionnaire and nurse interview. Health history (atrial fibrillation, rheumatoid arthritis, migraines, severe mental illness, erectile dysfunction) were extracted as the earliest event date from questionnaire, nurse interview, and linked health-records.

#### Follow-up and outcome definition

CVD was defined as a composite of fatal and non-fatal events to match the definition used to train and evaluate QRISK3. CVD events and dates were identified using existing codelists from self-reported questionnaire data, and extracted from linked ICD-9/10 and OPCS-4 in Hospital Episode Statistics (HES) records and death registries (see Table S4).[11] Participants were followed from baseline assessment visit until an incident CVD event, death, or 10 years of follow-up, whichever occurred first.

Participants were excluded if they withdrew consent, had prevalent CVD or lipid-lowering medication use at baseline (Table S5), were older than 70, or had missing key variables (BP, lipid measurements, IMD), yielding 327,923 individuals for analysis (Figure S1).

### 3.2 Genetic data and PRS

Genotyping and imputation along with QC of the UKB genetic data has been previously described.[37] To quantify genetic risk for CVD, we selected a well-studied PRS (metaGRS_CAD_; PGS000018)[9] with characteristic performance that has been extensively validated in external cohorts and diverse ancestries.[40, 41] We ensured no overlap between the PGS000018 training samples and our current analysis cohort of UKB, and identified three additional CAD PRS that did not inlcude UKB data in training as additional scores for sensitivity analyses (PGS000012,[8] PGS001780,[42] and PGS002244 [43]). These additional scores have broadly similar performance but differ in their construction: number of risk factors/SNPs; PRS training algorithms; different sized and ancestrally diverse development cohorts.

PRS were extracted from the Polygenic Score Catalog and calculated on UKB imputed genotype data using the *pgsc calc* tool, an established pipeline for reproducible PGS calculation and genetic similarity analysis.[44, 45] Genetic similarity to reference ancestry groups (superpopulations) was performed using the highest probability prediction based on comparison of PC loadings to the combined HGDP+1kGP population reference panel.[46] To ensure that PRS distributions were comparable between different ancestry populations, *pgsc calc* implements three different normalisation methods: most similar population (MSP), where a reference population that shares similar genetic ancestry with each individual was identified, and the individuals’ PRS was adjusted based on the distribution of PRS values within that reference group; normalised score 1 (N1), where the PRS distributions across ancestry groups were centred to mean 0 using a continuous representation of genetic ancestry (PCA space);[47] Normalised score 2 (N2), where the mean and variance was normalised across genetic ancestry groups using a continuous representation of genetic ancestry to have mean 0 and standard deviation 1.[18] All three normalisation methods were implemented for all PRSs (Figure S2). Percentiles of genetic risk used for reclassifying intermediate risk individuals were defined using the full subset of UK Biobank with genetics data (excluding metaGRS_CAD_ training samples) to ensure representativeness (n_genotyped_ = 487,180).

### 3.3 CVD risk prediction, recalibration, and reclassification based on PRS

The QRISK3 10-year CVD risk prediction model was calculated using the algorithm published by ClinRisk Ltd. at https://qrisk.org/src.php. Risk factors contributing to QRISK3 were: age, ethnicity, Townsend deprivation index, smoking status, SBP, standard deviation of SBP, height, weight, the ratio of total to HDL cholesterol, family history of myocardial infarction before the age of 60, atrial fibrillation, chronic kidney disease, erectile dysfunction, severe mental illness, migraines, rheumatoid arthritis, systemic lupus erythematosus, type 1 diabetes, type 2 diabetes, atypical antipsychotic medications, blood pressure treatment, and systemic corticosteroid usage (see Table S6). To ensure the baseline risk-prediction was optimised for the target population of UKB, we recalibrated the estimated QRISK3 predictions using a sex-specific rescaling method.[48] Sex-specific recalibration factors were computed for deciles of predicted risk by comparing observed 10-year Kaplan-Meier event rates with predicted probabilities (Table S7). Recalibration for age was not required as sex-recalibration resulted in acceptable calibration across age strata (Figure S3). Individuals were classified as low (*<*5%), medium (5-10%), or high ( ≥10%) risk based on UK national guidelines.[2] Individuals above the 10% threshold were classified as predicted to experience a CVD event in our one-stage risk straification.

To assess the utility of PRSs, a sequential two-stage model was implemented with PRSs as a risk-enhancing factor based on recommended implementation scenarios.[23] In the absence of standardised high-risk thresholds for PRSs, percentiles were used; existing implementation trials have used thresholds above the top 10% of the PRS distribution (selected to match the effect size of a positive family history).[17, 18] In this study, we tested thresholds of 1, 2, 5, 10, and 20%. Depending on their PRS and chosen threshold, intermediate-risk individuals were reclassified to the high-risk group based on the assumption that high PRS indicates elevated CVD risk (warranting earlier intervention). As recommended by guidelines, low-risk individuals were not up-classified due to other risk factors diminishing their overall risk.[22, 23] We evaluate the impact of varying PRS thresholds on test performance and fairness metrics. For each PRS threshold tested, we also compare to the results of up-classifying the same number of individuals based on the highest QRISK3 predictions in the intermediate risk stratum to test whether the changes in these metrics are specific to PRS rather than the expansion of the number of individuals classified as high-risk.

### 3.4 Fairness and performance metrics

We identified four measures of algorithmic fairness based on a scoping review of the literature, prioritising metrics that penalise false negative rates in order to maximise the number of at-risk individuals put forward for prevention (see A.1.1). These metrics require dividing the population into groups and calculating the difference between the predictions of each group, where the predictions are quantified using a selection of statistical measurements, which differ between metrics. Group fairness was assessed for both individual risk factors and PRS-integrated models using accuracy equality, equal opportunity, conditional use accuracy equality, and treatment equality (Table 1). Fairness disparities were quantified using the maximum pairwise group differences (Table S2): accuracy equality difference (AED), equal opportunity difference (EOD), conditional use accuracy equality difference (CUAED), and treatment equality difference (TED). The scale of the AED, EOD, and CUAED metrics is between 0 (fair across groups) and 1 (maximum unfairness).

**Table 1:** Fairness definitions for CVD risk prediction used in this analysis. Each definition is accompanied by a brief description and its corresponding mathematical formulation. The definitions focus on ensuring equitable model performance across demographic groups (*i, j* ∈ {*A, B*, …,*N*} ), addressing fairness from different perspectives through the use of different statistical measurements. To evaluate these fairness criteria we assess the maximum difference among groups of a focal characteristic (implementation described in Tables S2 and S3).

| Fairness definition | Definition description | Definition equation |
| --- | --- | --- |
| Equal accuracy | All groups have equal prediction accuracy (Acc). | $Acc_i = Acc_j$<br>$\forall i, j \in \{A, B, \dots, N\}, i \neq j$ |
| Equal opportunity [49, 50] | All groups have equal false negative error rates (FNR). | $FNR_i = FNR_j$<br>$\forall i, j \in \{A, B, \dots, N\}, i \neq j$ |
| Conditional use accuracy equality [30] | All groups have the same positive predicted value (PPV) and negative predictive value (NPV). | $PPV_i = PPV_j$<br>$\wedge NPV_i = NPV_j$<br>$\forall i, j \in \{A, B, \dots, N\}, i \neq j$ |
| Treatment equality [30] | All groups have the same ratio of false negative (FN) to false positive (FP) predictions. | $FN_i : FP_i = FN_j : FP_j$<br>$\forall i, j \in \{A, B, \dots, N\}, i \neq j$ |

To evaluate the performance of the binarised risk prediction model we choose the metrics of accuracy, sensitivity, specificity, precision. In the context of CVD risk prediction, sensitivity (recall) is of particular interest, as it is important to maximise the number of high risk cases detected in the form of true positives (which in turn minimises the false negatives).

### 3.5 Statistical analysis of fairness and performance metrics

The calculation of fairness and performance metrics is computed using a 2x2 confusion matrix, requiring binary risk predictions and a binary CVD outcome. Given the time-to-event nature of our 10-year CVD outcome data we account for 2.94% of individuals censored before 10 years (primarily non-CVD death or incomplete health record linkage) using inverse probability censoring weighting (IPCW). [51, 52] Kaplan-Meier estimates were used to compute IPCW weights:

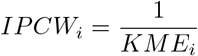

for non-events, CVD events were assigned the maximum weight. Individual’s weights were used to construct weighted confusion matrices to calculate fairness and performance metrics that downweight individuals who had less followup time. To estimate the variability of our results, we used bootstrap resampling (200 replicates) to estimate the 95% confidence intervals for all performance and fairness metrics.[53] As a sensitivity analysis, we performed 1000 replicates for the two-step analysis of PGS00018 to confirm that 200 replicates were sufficient to capture the variability observed in the fairness and performance metrics.

#### Individual risk factor analysis

To evaluate the fairness of individual risk factors as standalone predictors (SBP, non-HDL, T.Chol:HDL ratio, age, PRS) we varied the percentage of the population classified at high-risk using the top 1%, 2%, 5%, 10%, or 20% of the distribution. In addition, we added any clinical thresholds used to interpret SBP (*>*140) [54], non-HDL (*>*4), the T.Chol/HDL ratio (*>*6) [55], and suggested ages to initiate risk assessment (50, 55, and 60) [56]. To eliminate confounding and enable individual risk factors to be evaluated accurately, demographic differences across subgroup comparisons were balanced using a 1:1 nearest neighbour matching method (without replacement, similar to propensity score matching [57]) using a Euclidean distance metric. For each group within a characteristic, a “1:1 matched” population was created in each bootstrap replicate. This enabled fair comparison between matched groups for individual predictors (details in Supplementary Methods A.1.2). Fairness metrics are quantified as the maximum difference between each subgroup and the balanced comparison group (Table S3).

#### Two-step risk prediction analysis

To evaluate the utility of PRS, we varied the number of individuals predicted as high risk by reclassifying the top 1%, 2%, 5%, 10%, or 20% of the score distribution in the intermediate risk category to high risk. To analyse the overall predictive ability of the reclassification we used the IPCW-weighted confusion matrix to calculate accuracy, precision, sensitivity, and specificity of the original QRISK3 high-risk classification, the PRS-based reclassifications at varying thresholds, and the comparison group of adding the same number of individuals to the high-risk group as PRS using QRISK3. Change in performance was expressed as the absolute difference in the performance metric between the two-stage model (PRS or an equal number of people reclassified using QRISK3) and the one-stage model as baseline. The fairness metrics were also calculated on the two sets of comparisons, using the maximum difference between groups of a focal characteristic (Table S2). Change in fairness was expressed as the absolute difference in the fairness metric between the two-stage model and the one-stage model as baseline.

## 4 Results

### 4.1 Study Population

The study included 327,923 individuals (57.23% female; mean age 55.9 *±* 8.0 years) without a history of CVD or lipid-lowering medication use at baseline (Table 2, Figure S1). These data contain sufficient size and diversity to evaluate group fairness for the characteristics of interest (age, sex, self-reported ethnicity, IMD), despite being majority White ethnicity (92.3%). Some intersectional differences were observed among these demographic characteristics, particularly between ethnicity groups. Black ethnicity participants were disproportionately represented in the most deprived quintile (IMD Q5), with over half (57.68%) falling within this group. Asian and other ethnic groups also had a relatively high proportion of individuals in IMD Q5, while White ethnicity participants were more evenly distributed across all quintiles, with only 17.3% in IMD Q5. White participants were on average older compared to participants of other ethnicities. Among the entire cohort 5.67% of individuals experienced a CVD event in the 10-year followup (18,581 events).

**Table 2:**
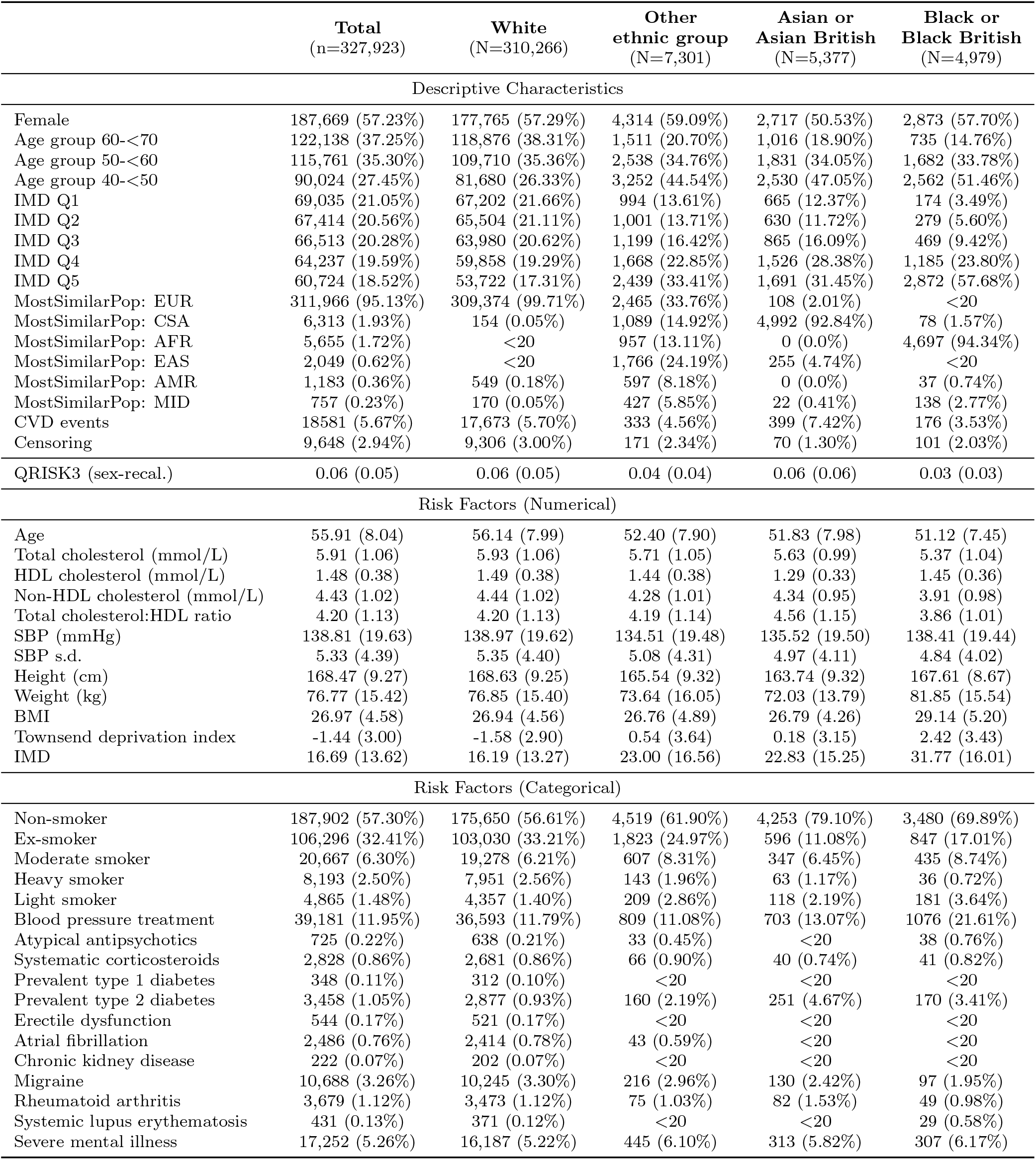
Descriptive characteristics of study cohort and CVD risk factors. Stratification by self-reported ethnicity. Counts with greater than 20 participants (and %) are provided for categorical variables. Means (and s.d.) are provided for continuous variables. *pgsc calc* was used to define the most similar superpopulation (MostSimilarPop) label for each sample in the HGDP+1kGP reference panel.

### 4.2 Evaluating the fairness of individual risk factors for CVD risk-stratification

Individual risk factor measurements and clinically-defined thresholds are often used to identify individuals at high-risk of CVD. While these thresholds and tests may have known performance characteristics in relevant subgroups (e.g. sensitivity, specificity), their fairness characteristics among key demographics are underexplored. To measure the fairness of each risk factor, we defined varying percentiles of the risk factor distribution as high-risk (including clinically relevant thresholds) and evaluated the group fairness metrics for each threshold. To ensure that the fairness measures for specific demographics are not confounded by intersectional differences among other focal characteristics, we ensured that comparisons were balanced using a 1:1 matching and bootstrapping approach (see 3.1 and A.1). This matching procedure yields balanced comparisons (Figure S4), with only minor deviations in ethnicity distributions among the age, sex, and deprivation subgroups (mean Euclidean distance in ethnicity distributions *<* 0.02) .

Across all risk factors and demographic groups, fairness metric values were low (typically below 0.2), except for TED which may exceed 1 due to its ratio-based definition (Figures 1, 2, 3, 4). In general, these results suggest limited disparity across most single risk factors and thresholds. Across all thresholds, fairness was lowest and most stable for IMD and age groups. Ethnicity groups exhibited the greatest variation in fairness, particularly at extreme thresholds and across PRS normalisation methods.

**Figure 1:**
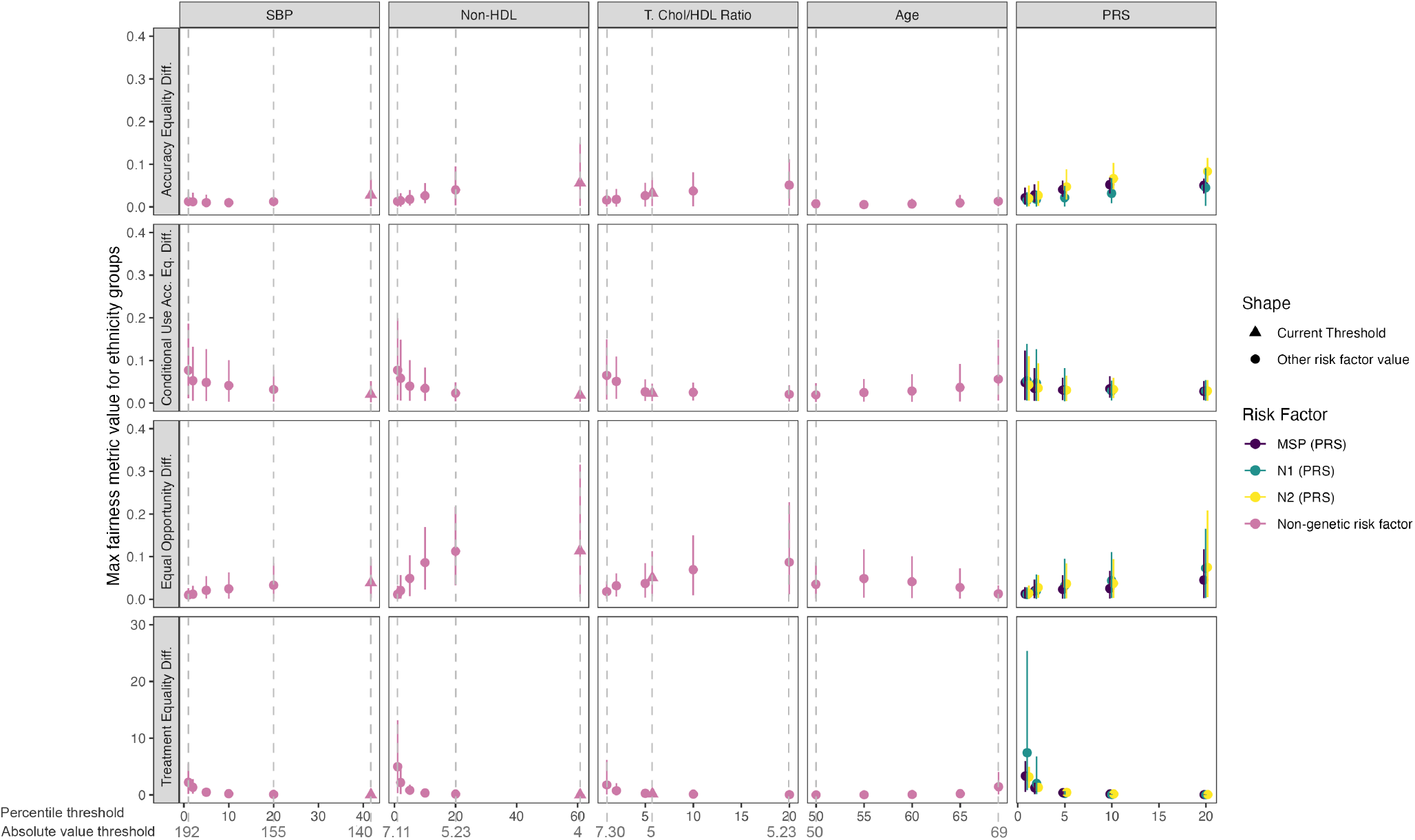
Fairness metrics for ethnicity matched groups. Mean fairness values for accuracy equality difference (AED), conditional use accuracy equality difference (CUAED), equal opportunity difference (EOD), and treatment equality difference are presented across matched ethnicity groups for all risk factors (SBP, non-HDL cholesterol, total cholesterol:HDL, age, and PRS). Error bars refer to the 95% confidence intervals calculated across 200 bootstraps replicates. Increasing values indicate increasing unfairness, the maximum possible value for AED, CUAED, and EOD is 1. Each panel corresponds to a specific risk factor, with shapes distinguishing current clinical thresholds (triangles) from other evaluated thresholds (circles). PRS normalisation approaches (MSP, N1, and N2) are displayed in different colours. Percentile and absolute value thresholds are indicated along the x-axis.

**Figure 2:**
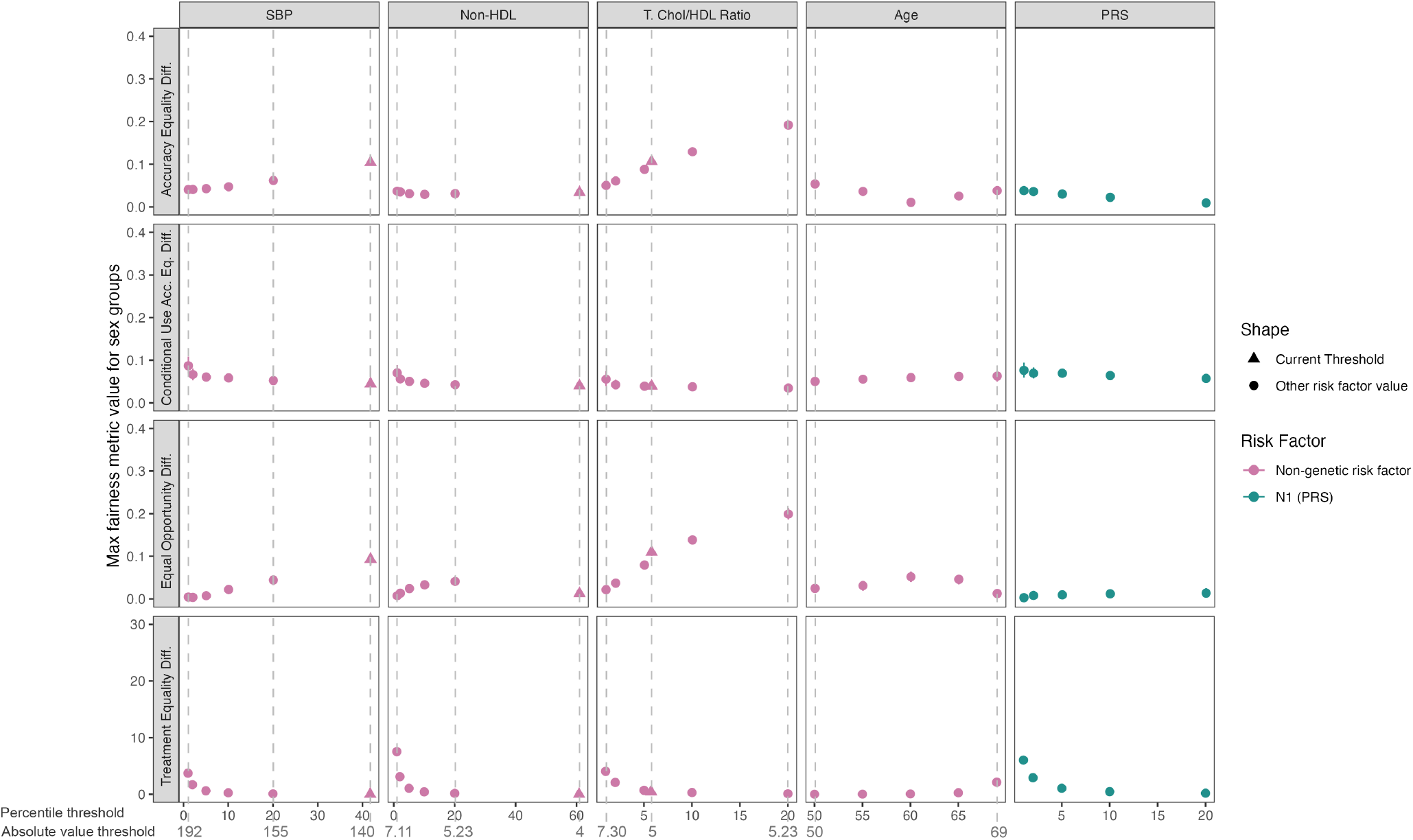
Fairness metrics for matched sexes. Mean fairness values for accuracy equality difference (AED), conditional use accuracy equality difference (CUAED), equal opportunity difference (EOD), and treatment equality difference are presented across matched sexes groups for all risk factors (SBP, non-HDL cholesterol, total cholesterol:HDL, age, and PRS). Error bars refer to the 95% confidence intervals calculated across 200 bootstraps replicates. Increasing values indicate increasing unfairness, the maximum possible value for AED, CUAED, and EOD is 1. Each panel corresponds to a specific risk factor, with shapes distinguishing current clinical thresholds (triangles) from other evaluated thresholds (circles). Percentile and absolute value thresholds are indicated along the x-axis.

**Figure 3:**
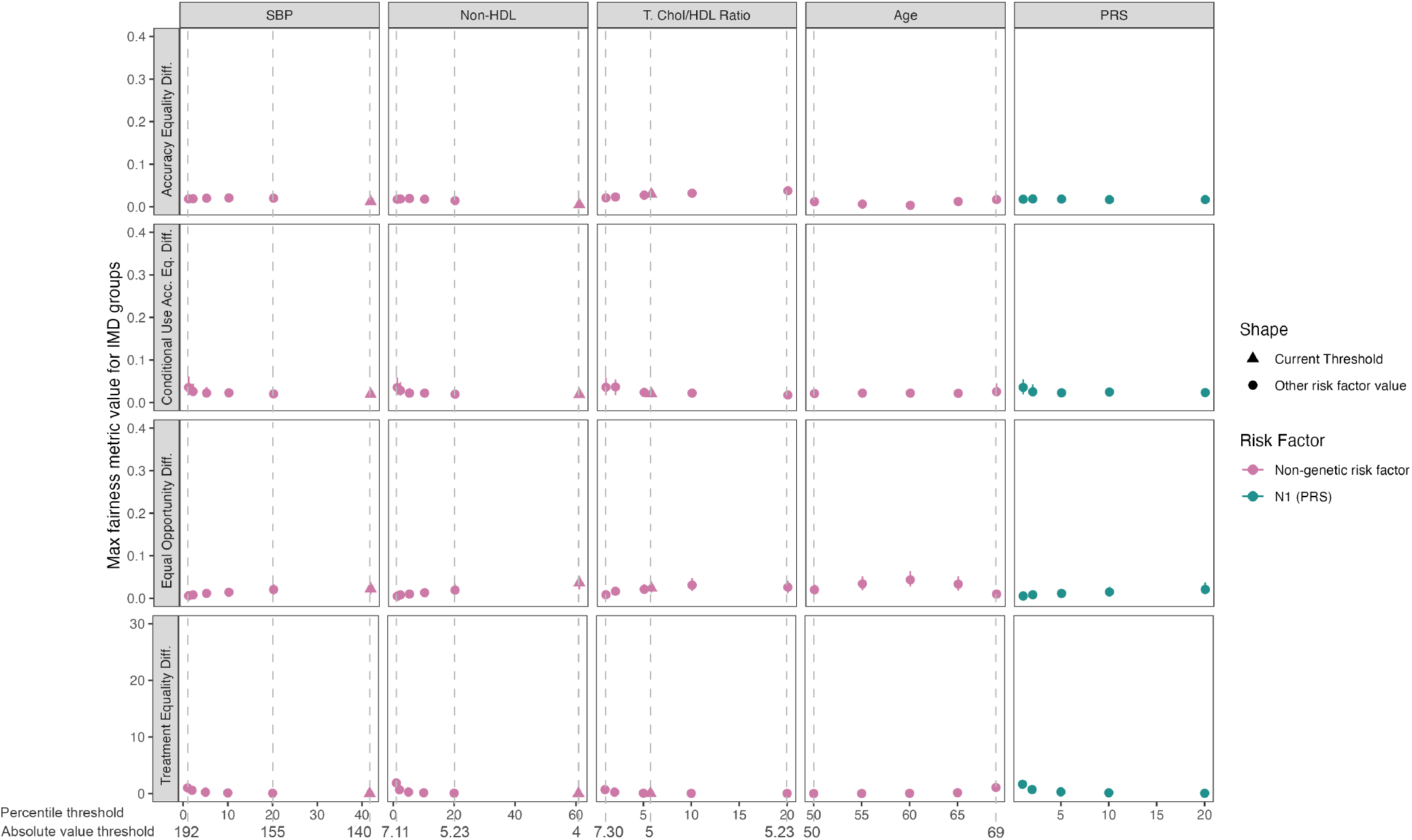
Fairness metrics for IMD matched groups. Mean fairness values for accuracy equality difference (AED), conditional use accuracy equality difference (CUAED), equal opportunity difference (EOD), and treatment equality difference are presented across matched IMD groups for all risk factors (SBP, non-HDL cholesterol, total cholesterol:HDL, age, and PRS). Error bars refer to the 95% confidence intervals calculated across 200 bootstraps replicates. Increasing values indicate increasing unfairness, the maximum possible value for AED, CUAED, and EOD is 1. Each panel corresponds to a specific risk factor, with shapes distinguishing current clinical thresholds (triangles) from other evaluated thresholds (circles). Percentile and absolute value thresholds are indicated along the x-axis.

**Figure 4:**
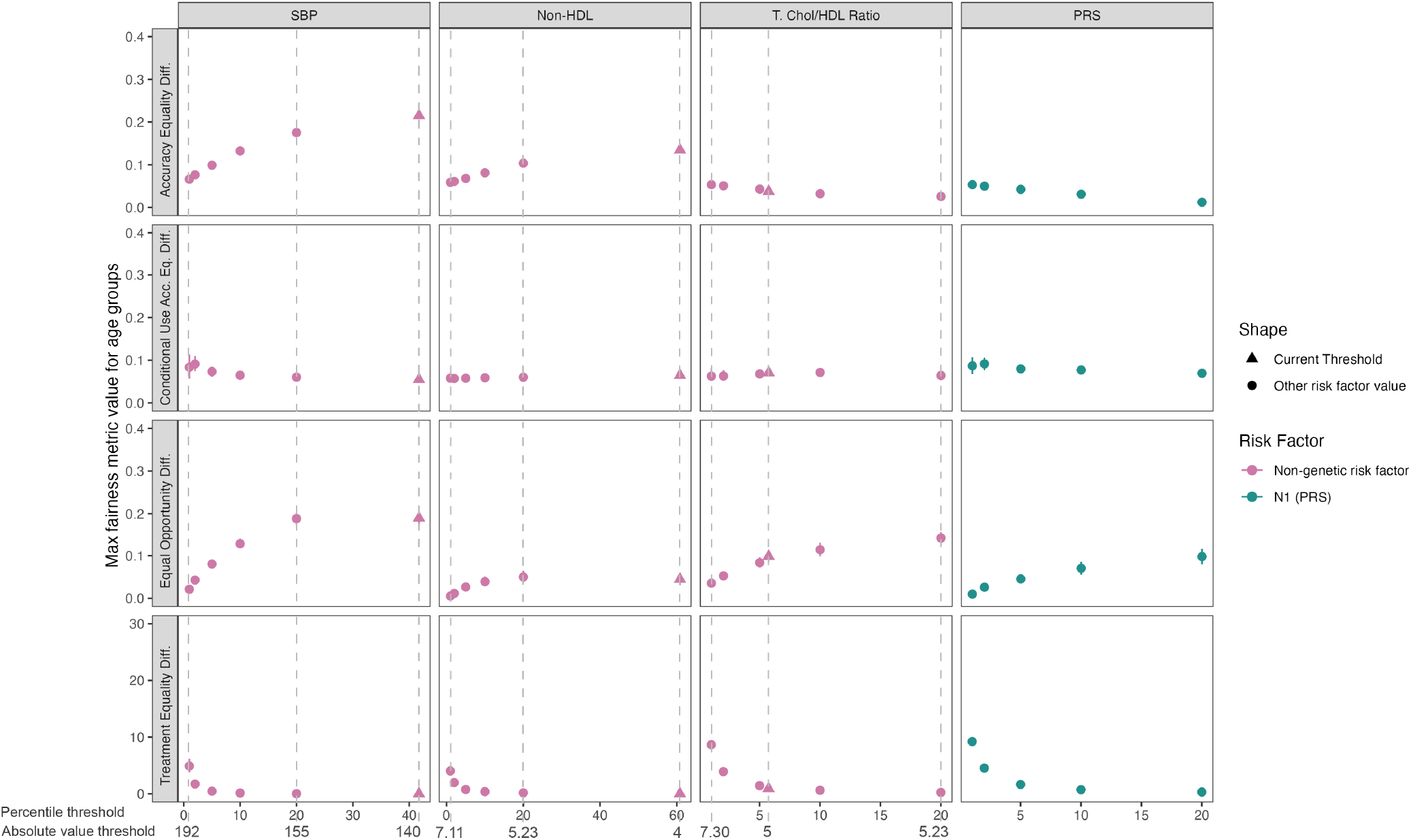
Fairness metrics for age matched groups. Mean fairness values for accuracy equality difference (AED), conditional use accuracy equality difference (CUAED), equal opportunity difference (EOD), and treatment equality difference are presented across matched age groups for all risk factors (SBP, non-HDL cholesterol, total cholesterol:HDL, age, and PRS). Error bars refer to the 95% confidence intervals calculated across 200 bootstraps replicates. Increasing values indicate increasing unfairness, the maximum possible value for AED, CUAED, and EOD is 1. Each panel corresponds to a specific risk factor, with shapes distinguishing current clinical thresholds (triangles) from other evaluated thresholds (circles). Percentile and absolute value thresholds are indicated along the x-axis.

For the PRS, three different methods (MSP, N1, N2) were applied to normalise the differences in score distri-butions that occur across ancestry groups, resulting in relative genetic risk estimates that are comparable across individuals and defined with respect to the entire UKB population (Figure S2). The different normalisation methods showed similar trends within each evaluated fairness metric between ethnicity groups and across different thresholds of the PRS distribution used to define high-risk (Figure 1). At less stringent thresholds (top 20% of PRS), the N2 method had the highest levels of unfairness in AED (0.08, 95% CI=[0.06, 0.12]) and EOD (0.08, 95% CI=[0.01, 0.21]), indicating larger maximum differences in accuracy and FNRs across ethnicity groups. At the highest levels of genetic risk (top 1% of PRS), the N1 method was observed to have higher unfairness in TED (7.42, 95% CI =[0.86, 25.38]). Generally, the trends in fairness metric changes across percentiles were less apparent for the MSP method, which also had lower values than the conventional single risk factors. The trends observed for fairness of PRS and different normalisation methods among ethnicities were generally consistent with three other comparable scores used as sensitivity analyses (Figure S5). For comparisons of PRS fairness among sex, age, and IMD groups, the choice of normalisation method or PRS did not affect the results (Figures S6, S7, S8). Given that the advantages of the N1 method (e.g. that it does not require explicit comparison with reference populations) and its equivalence to MSP at thresholds that have been used to define high-risk in clinical trials (e.g. top 10%), we chose to use the results for the N1 normalisation method as exemplar and treat the other methods as sensitivity analyses in subsequent analyses.

Although the unfairness of CVD risk factors between sex groups was low overall (*<* 0.2), there exist some interesting fairness patterns in SBP and the two different non-HDL cholesterol measurements (Figure 2). For SBP a threshold of 140 mmHg increases unfairness in AED and EOD, likely because it classifies ∼40% of the population as high-risk. Interestingly, the two measures of lipid risk differ significantly in AED patterns between sexes (Figure 2), and the opposite pattern exists when comparing different age groups (Figure 4).

When considering different age thresholds as a risk factor for CVD (e.g. age to initiate risk assessments) we observed typically flat levels of unfairness among ethnicity, sex, and deprivation groups (Figures 1, 2, 3). The lowest age threshold evaluated (50), exhibited less unfairness compared to older age thresholds across all metrics and characteristics. Some disparities were observed when comparing the fairness of all other risk factors across age groups (Figure 4). The highest values across the age group analysis were observed for SBP in AED and EOD, which indicated that the greatest observed differences were observed in accuracy and FNRs for SBP rather than other risk factors. The increase in fairness metric values (indicating greater unfairness) as the threshold increased were also observed for non-HDL in AED, and for total cholesterol:HDL in EOD. The remaining risk factors’ metric values were generally low with minimal difference between thresholds, and there were slightly larger values for TED at the low percentile thresholds. Across age groups, the metrics’ values were higher than for most of the other characteristics (for SBP, non-HDL and total cholesterol:HDL but not PRS).

### 4.3 Evaluating the impact of PRS as a risk-enhancing factor for population risk-stratification

Current research and statements from clinical groups suggest that PRS for CVD can serve as risk-enhancing factors, particularly among individuals predicted to be intermediate risk using established risk calculators.[10, 15, 22, 23] To measure the impact of PRS on population-level risk-stratification, we first calculated QRISK3 (UK guideline-recommended CVD risk calculator) and applied sex-based recalibration to ensure accurate predictions in our cohort (Figure S3). The mean predicted 10-year CVD risk in our recalibrated model is 6% (Table 2; guideline-established risk thresholds stratify the population into 181,215 low (*<* 5%) risk, 89,145 intermediate (5-10%) risk, and 57,563 high (*>* 10%) risk individuals (Figure 5A).

**Figure 5:**
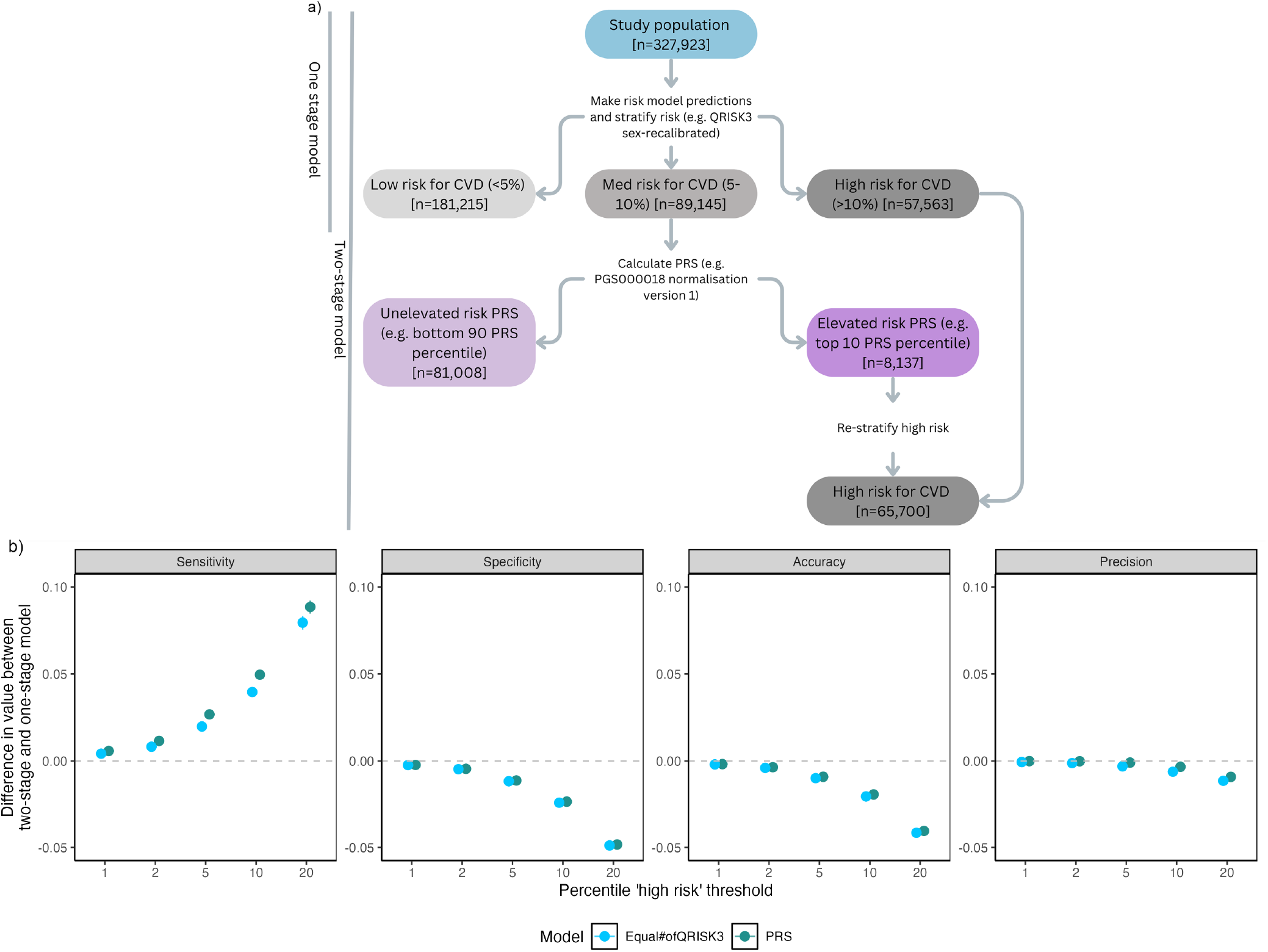
Impact of two-stage risk stratification including PRS on risk-stratification performance. (a) Current CVD risk assessments involve a single risk prediction (recalibrated QRISK3), stratification into low, intermediate, and high-risk is based on guideline-recommended treatment thresholds. To evaluate the use of PRS as a risk-enhancing factor (two-stage model), the PRS is applied to intermediate risk individuals to reclassify individuals as high-risk (numbers correspond to the reclassification of top 10% of risk using PGS000018 N1). (b) Differences in performance between the two-stage and one-stage model are displayed for sensitivity, specificity, accuracy and precision (Table S10). Mean difference (one-stage model as baseline) are calculated for varying PRS thresholds (PGS000018 N1), error bars correspond to 95% confidence intervals across 200 bootstraps.

When applying PRS as a risk-enhancing factor to the intermediate-risk cohort, individuals in the top percentiles of the PRS distribution were reclassified into the high-risk group. For example, using the top 10% of PRS (N1) distribution an additional 8,137 individuals re-classified as high-risk resulting in 65,700 individuals (Figure 5A). The number reclassified varied by PRS threshold and normalisation method (Supplementary Figure S9). Overall, up-classified individuals were typically from risk-enhancing groups (older males of White ethnicity), reflecting the existing high-risk classifications of the baseline risk prediction.

Next, we measured the impact of PRS-based reclassification (two-step model) in comparison to using QRISK3 alone (one-stage model) on test performance metrics in the entire cohort. Using PRS to reclassify intermediate-risk individuals into the high-risk category improves sensitivity (TPR/recall) without significant compromises in precision (Figure 5B; Supplementary Table S10), with very little variation across bootstrapped replicates. All thresholds of PRS used to define high-risk showed an increase in sensitivity - for example, using the top 10% of PRS, increased sensitivity by 5.0% (95% CI: [4.7, 5.2]), equating to 924 additional CVD cases identified (compared to 739 when expanding the high risk set with QRISK3). At the same 10% threshold, specificity and accuracy declined slightly by 2.3% and 1.9%. Nearly identical performance metrics were observed for the three other PRS for CVD (Figure S11), which is expected given the similar scale of effect sizes for population-level risk stratification (Figure S2) that has also been previously observed.[58] Identifying additional high-risk individuals using PRS consistently outperforms reclassifying the same number of individuals by lowering the QRISK3 threshold, with greater sensitivity increases and slightly smaller declines in specificity, precision, and accuracy (Figure 5). Overall, the performance metrics illustrate that applying PRS in a two-step risk-stratification identifies additional high-risk individuals that develop CVD over the 10-year follow-up period.

### 4.4 Evaluating the fairness of including PRS in two-step CVD risk-prediction

Many existing clinical risk prediction models are known to have sub-optimal fairness.[59–63] When evaluated using our approach, the recalibrated QRISK3 model alone displays notable levels of unfairness for all characteristics other than IMD quantified by: EOD (up to 0.44 [0.30, 0.61] for ethnicity); AED (up to 0.35 [0.34, 0.35] for age); and TED (up to 3.46 [3.15, 3.83] for age) (Table S9).

Given the well-documented issues regarding the transferability of PRS between ancestry groups and subpopulations,[25] and the low but detectable levels of unfairness detected using PRS as a standalone risk-factor (Figure 1), we next sought to evaluate if including PRS as a risk-enhancing factor changes the overall fairness of risk prediction (Figure 6). Across sex, age, and IMD groups, changes in fairness were minimal for PRS as a risk-enhancing factor. The largest observed value was for EOD at 0.079 (95% CI: [0.033, 0.12]) for fairness among ethnicity groups (top 20% of PRS reclassified). The largest decreases were seen for TED between age groups at the 20th percentile threshold with an improvement of -2.53 (95% CI: [-2.81, -2.26]).

**Figure 6:**
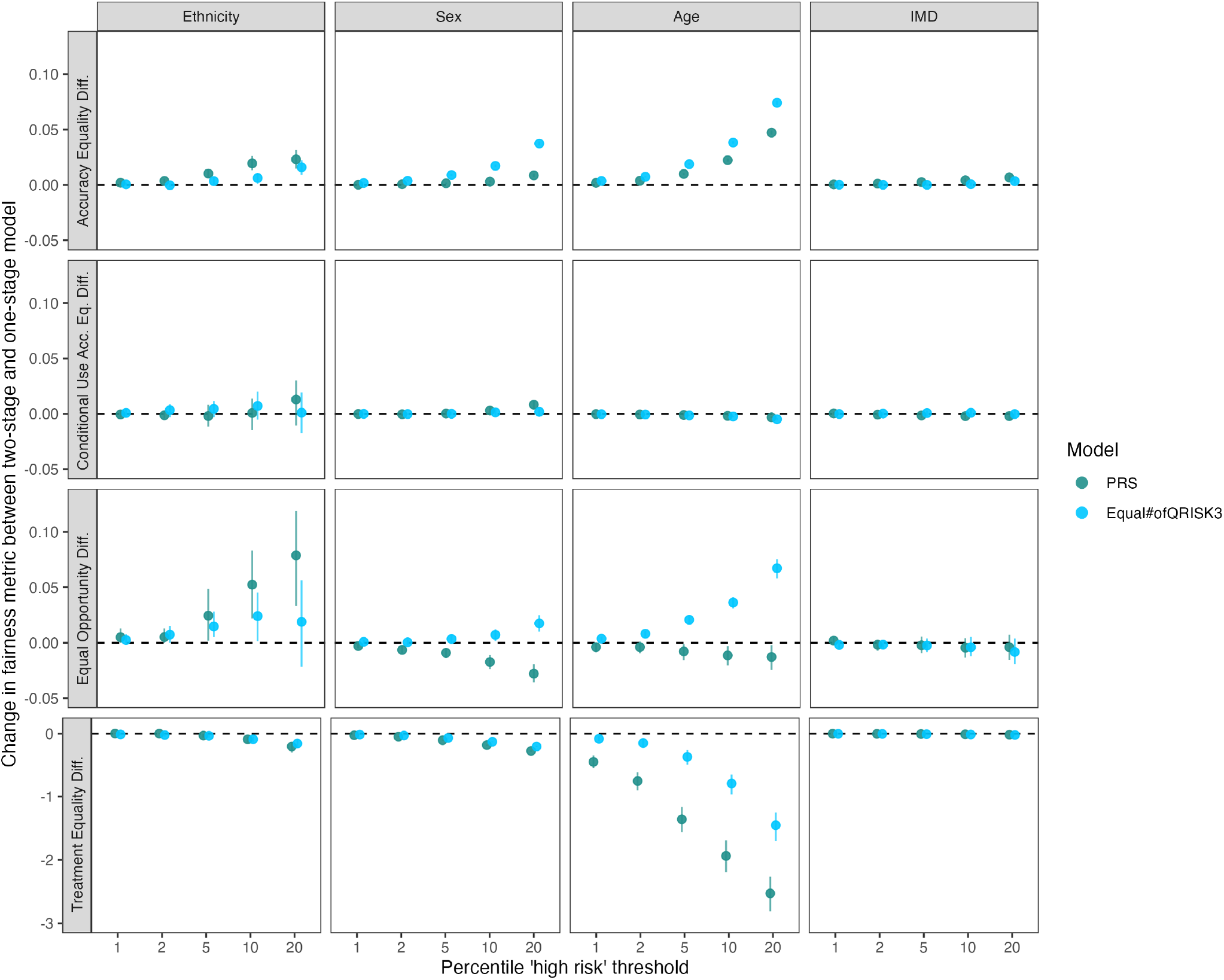
Difference in fairness metric values between two-stage (PRS-added) and one-stage risk predictions. Changes in metrics for ethnicity, sex, age, and IMD characteristics were evaluated for the QRISK3 sex-recalibrated baseline model and their two-stage PRS-added versions (PGS000018 N1). Mean difference in values for accuracy equality difference, conditional use accuracy equality difference, equal opportunity difference, and treatment equality difference between two-stage and one-stage models. Error bars correspond to 95% confidence intervals calculated across 200 bootstrap replicates. Positive values indicate increasing unfairness compared to the baseline QRISK3 model, negative values indicate fairness improvements.

The impact of PRS on fairness across ethnicity groups observed in the standalone risk factor analysis (Figure 1) was also observed in the two-stage analysis (Figure 6). At the 10% PRS threshold, small absolute differences in AED (0.019, 95% CI: [0.013, 0.026]) and EOD (0.052, 95% CI: [0.022, 0.083]) were observed; however, this was coupled with a small benefit in TED (-0.089, 95% CI: [-0.142, -0.047]). Interestingly, at the same threshold the comparison reclassification using QRISK3 resulted in either larger unfairness or smaller benefits among age and sex groups, which were particularly pronounced for EOD. Our results using PGS000018 were generally consistent across PRS normalisations (Figure S12) and alternative PRS (Figure S13), with similar fairness metrics across all characteristics. The robustness of the fairness results across PRS and normalisation methods is expected based on the correlation and similar effect sizes between the predictors (Figure S2B-C).

To ensure that our results were not impacted by the choice of risk thresholds or recalibration methods, we performed a sensitivity analysis where we varied the size and thresholds used to define high risk, varying cutpoints between 5 and 10% risk (Figures S14, S15). The potential for the PRS to improve sensitivity increases with the threshold used to define high risk, and among the cutoffs where the percentage of the population defined as intermediate risk is greater. PRS outperforms traditional risk factors except for a few scenarios where a very strict risk threshold (20%) is used to define high-risk. The fairness patterns in EOD and AED are also similar among all definitions of intermediate risk (including our main 5-10% threshold) -the few scenarios where using QRISK3 does outperform PRS are coupled to increases in age and sex unfairness. Overall, the two-stage model including PRS meaningfully increases specificity and is coupled to smaller fairness changes with some benefits over using traditional risk factors alone.

## 5 Discussion

In this study, we evaluated the equitable performance of CVD risk factors, PRS, and risk prediction models using algorithmic fairness metrics relevant to clinical risk prediction. The fairness metrics were selected to quantify parity in false negative rates to ensure that individuals who do experience CVD events are labelled as high risk, where they will have an equal opportunity for safe and effective treatments (e.g. statins). We applied these metrics to measure fairness across key protected characteristics (age, sex, ethnicity) and measures of potential health-inequity (IMD) available in UKB. When evaluating individual CVD risk factors using the group fairness measures, we observed low levels of unfairness across all measured characteristics ( *<* 0.2 for AED, CUAED, and EOD metrics). When varying the thresholds used to classify individuals at high risk, the AED and EOD fairness metrics typically increase as a larger proportion of the population is classified, while CUAED and TED tend to decrease. This pattern reflects how fairness metrics respond differently to the sensitivity-specificity trade-off, and can be used to define optimal thresholds. Examining the risk factor fairness patterns across all characteristics evaluated in this study identified unique associations, including PRS with ethnicity groups, measures of non-HDL cholesterol with sex, and SBP with age. When PRS were incorporated as risk-enhancing factors into existing risk prediction frameworks, we observed increases in specificity and minimal differences in fairness metrics, indicating that PRS are unlikely to meaningfully exacerbate disparities.

The concern that PRSs may exacerbate health inequities due to known limitations in transferability between ancestry groups has garnered increasing attention in recent years.[28, 29] It is well documented that the best PRS for CAD have inconsistent effect sizes in different ancestry groups [41, 64, 65]; however, we and others have argued that PRS may still be useful to identify high-risk individuals who may benefit from treatment in all ancestry groups.[6] When using PRS alone to define high-risk individuals, the highest unfairness was observed among ethnicity groups; however, the unfairness of PRS in these groups was not significantly different from the fairness observed for guideline-recommended thresholds for SBP and non-HDL cholesterol. Variable transferability of PRS across ancestry groups likely causes the observed differences in ethnicity group fairness given their overlapping distributions in our cohort, but differences between ethnicity groups also capture other factors associated with social determinants of health.[66] In our analysis, the three most common methods used to account for genetic ancestry in estimating relative genetic risk using PRS also differentially impact fairness across ethnicity groups and not the other characteristics. PRS risk classified using the N2 method of normalisation results in higher AED and EOD unfairness when larger proportions of the population are selected (*>*10%), whereas the N1 and N2 methods have more variable performance in CUAED and TED at stringent thresholds (*<*5%). All PRS constructions and normalisations defined a similar number of individuals as high-risk based on population percentiles; however, the incidence of CVD between ethnicity groups is variable, and these absolute risk differences cannot be captured by current PRS. These results confirm that PRS are less likely to be useful as stand-alone risk predictors, and are more useful when combined with additional risk factors to predict absolute risk.

Unique fairness patterns were also observed for clinical risk factors, which can also be interpreted using guideline-recommended thresholds to define high-risk individuals (unlike the distribution of PRS which has no established cutpoints). Particularly interesting is that the two different cholesterol measurements used to define high-risk of CVD in the clinic behave differently in fairness between sexes, particularly for the T.Chol/HDL ratio across metrics and thresholds. Given the age distribution in our cohort, this pattern may be driven by the change in blood cholesterol profiles that occurs during menopause.[67, 68] Across all tested characteristics it is clear that the recommended risk thresholds for HDL cholesterol and SBP were not representative of actual the ‘high risk’ CVD population as they captured a large proportion of individuals in our cohort( ∼ 40 and ∼ 60% respectively) yielding unfairness among sexes and age groups. Although not accurate on their own, these measurements are likely to be used as a liberal cut-off for prioritising individuals who require investigation using an integrated risk calculation that can be used to define preventative treatment decisions, here the UK-guideline recommended tool (QRISK3).

Previous analyses of the QRISK3 model have shown that it has moderate calibration overall in UK Biobank, but poor calibration in specific subgroups such as older people.[69] Previous work has also stressed the importance of analysing models beyond calibration [70], for example by investigating inequities in subgroups. By using algorithmic fairness metrics here, we gain insight into the types of prediction differences and potential shortcomings of the model. In our cohort, fairness analysis in QRISK3 alone revealed varying levels of unfairness across different metrics and demographic groups (Table S11), representing its first evaluation using algorithmic fairness metrics in UKB. From the perspective of the evaluated characteristics, fairness varied depending on the metric used, highlighting the importance of exploring multiple metrics (despite most analysis focusing on a single metric).[24, 71] Fairness differences may indicate that appropriate stratification of baseline risks or interaction terms in the prediction model are either not included or not sufficiently capturing the different relationship of a risk factor across subgroups - incorporating fairness evaluation during model construction may help to improve future risk predictions. Exploring multiple fairness interpretations allows one to observe model behaviour in a more nuanced way, highlighting trade-offs and investigating performance differences across population groups; hence providing a more thorough approach than traditional discriminative measures.

In this study we evaluated the use of PRS as a risk-enhancing factor in intermediate-risk individuals, a scenario that has been evaluated in research studies and recommended by professional societies like the American Heart Association and European Society of Cardiology.[22, 23] Our results support the potential utility of PRS in this use case. All thresholds of genetic risk used to reclassify individuals as high-risk improved sensitivity - identifying more individuals who subsequently developed CVD - while maintaining comparable precision. The sensitivity gain was also specific to the use of PRS, reclassifying an identical number of people using traditional risk factors does not carry the same benefit. Importantly, the improvement in case detection did not come at the expense of fairness, differences in AED and CUAED were minimal among all characteristics tested. The addition of PRS slightly increases unfairness in EOD among ethnicity, age, and sex groups but improves the fairness using the TED metric - the addition of PRS makes the FNRs slightly different between groups while making the FNR:FPR ratio more balanced. On the absolute scale, the effect on EOD (*<*0.1) is very small and unlikely to result in harm at more stringent thresholds for reclassifying individuals as high-risk (e.g. 10%). As we also show in the standalone risk factor analysis, existing risk factors also carry some ethnicity biases, and incorporating PRS into risk stratification is less unfair by age and sex compared to using QRISK3 alone. These fairness differences partially stem from the different CVD incidences among age, sex, and ethnicity groups, the application of a second integrated risk calculator using PRS or different PRS thresholds may correct for these issues.[10] In sensitivity analyses, our results are robust across different PRS constructions. While the scores may rank different individuals at high-risk, their consistency in population performance to identify additional incident CVD-events is expected.[58] To our knowledge, these results represent the first application of algorithmic fairness metrics to evaluate the inclusion of a novel risk factor in a risk-stratification framework, and underscore the potential of PRS to enhance early detection of CVD risk in a clinically meaningful and equitable manner at the population-level.

Our study confirms previous results illustrating the added benefit of using PRS to improve CVD risk-stratification of individuals likely to benefit from preventative treatment, and applies algorithmic fairness metrics to suggest that PRS will not significantly increase any inequity; however, several limitations merit consideration. First, although we examined fairness across multiple dimensions (age, sex, ethnicity, IMD), these characteristics are unlikely to capture the full complexity of social determinants of health, specifically intersectional or context-specific sources of inequity.[72, 73] Second, although all fairness metrics were selected to be relevant to CVD risk prediction (to prescibed safe and low-cost treatments like statins), they may not be direcltly relevant to other medicines or conditions and different metrics can provide conflicting results: it is well-described that not all fairness criteria can be optimally satisfied.[74–77] While the metrics themselves do not readily allow the selection of a single best cutoff, the plots allow for the visualisation of unfairness that may not be readily apparent when using a single metric. Third, although we used multiple validated PRSs and normalisation strategies, all PRSs in this study were derived from cohorts with predominantly European ancestry, which may overestimate the unfairness of PRS compared to scores developed using better methods and more diverse training data. Finally, our analysis was conducted in UKB, a large prospective cohort with a comprehensive set of exposures and linked health records to assess outcomes; however, the limited sample size for minority ethnic participants and known healthy volunteer bias of the cohort may result in underestimation of unfairness compared to real-world settings.[78] Based on the assumption that risk factor associations discovered in UKB are broadly representative of the wider population,[79] and our methods to correct for differences between subgroups using matched populations, we assume that our fairness evaluations are representative for at least the portion of the UK population overlapping the UKB population. The availability of larger, more representative cohorts like Our Future Health with linked genomic and health-data in conjunction with the planned release of nationally-representative sampling weights will allow for replication of these findings once significant follow-up has accrued.

In conclusion, our results provide empirical evidence that PRS, when incorporated into integrated CVD risk assessments, are unlikely to meaningfully exacerbate existing disparities across key demographic groups. This supports current clinical recommendations for the use of PRSs as risk-enhancing factors and suggests that their correct application can improve population-level risk stratification while maintaining fairness.

## 6 Data Availability

Individual-level data from the UK Biobank is available to bona fide researchers, access can be requested by registering and submitting a project application (https://www.ukbiobank.ac.uk/).

## 7 Acknowledgements

This research has been conducted using the UK Biobank Resource under Applications 608471 and 7439, linked health data was provided by patients and collected by the NHS as part of their care and support. CC was supported by a Wellcome Trust and Health Data Research UK (HDR-UK) PhD studentship (PHD2020CAM003). SCR and LP were supported by British Heart Foundation Cambridge Centre of Research Excellence (CRE) Career Development Fellowships (RE/24/130011). AMW was supported by BHF Data Science Centre (HDRUK2023.0239), HDR-UK (Big Data for Complex Disease-HDR-23012), and as an NIHR Research Professor (NIHR303137)[*]. M.I. was supported by the Munz Chair of Cardiovascular Prediction and Prevention as well as the UK Economic and Social Research 878 Council (ES/T013192/1). This work was also supported by core funding from the British Heart Foundation (RG/18/13/33946; RG/F/23/110103), NIHR Cambridge Biomedical Research Centre (NIHR203312) [*], BHF Chair Award (CH/12/2/29428), Cambridge BHF Centre of Research Excellence (RE/18/1/34212), and by HDR-UK, which is funded by the UK Medical Research Council, Engineering and Physical Sciences Research Council, Economic and Social Research Council, Department of Health and Social Care (England), Chief Scientist Office of the Scottish Government Health and Social Care Directorates, Health and Social Care Research and Development Division (Welsh Government), Public Health Agency (Northern Ireland), British Heart Foundation and the Wellcome Trust. This work was performed using resources provided by the Cambridge Service for Data-Driven Discovery (CSD3) operated by the University of Cambridge Research Computing Service (www.csd3.cam.ac.uk), provided by Dell EMC and Intel using Tier-2 funding from the Engineering and Physical Sciences Research Council (capital grant EP/P020259/1), and DiRAC funding from the Science and Technology Facilities Council (www.dirac.ac.uk).

*The views expressed are those of the authors and not necessarily those of the NIHR or the Department of Health and Social Care.

