## Supplementary Text, Figures, and Tables for "Current polygenic risk scores are unlikely to exacerbate unfairness in cardiovascular disease risk prediction"

### A Supplementary Material

#### A.1 Supplementary methods

##### A.1.1 Fairness Metric Selection Rationale

A scoping review of the algorithmic fairness literature was undertaken to inform the selection of fairness definitions most suitable for clinical risk prediction. This process identified numerous fairness metrics, many with context-specific assumptions. Table S1 summarises key definitions, classified by group, subgroup, or individual fairness, following the taxonomy in [1]. Definitions marked with an asterisk (*) were shortlisted for analysis in this study.

Table S1: **Prominent fairness definitions and their grouping types.**

| Fairness definition name(s) | Grouping type |
|---|---|
| Equal Accuracy* | Group |
| Equal Opportunity* [2]/False Negative Error Rate Balance [3] | Group |
| Equalised Odds* [2]/Error Rate Balance /Separation [3–5] | Group |
| Demographic Parity [6–9]/Interdependence/Statistical Parity | Group |
| Predictive Parity/False Positive Error Rate Balance/ Predictive equality* [3] | Group |
| Unconstrained Utility Maximisation with Single-Threshold [10] | Group |
| Sufficiency [11] | Group |
| Ratio of False Negatives to False Positives [12] | Group |
| Avoiding Disparate Mistreatment [13] | Group |
| AUC Parity/Equality of Accuracy [14] | Group |
| Calibration within Groups [15]/Well-Calibration [16] | Group |
| Calibration between Groups [17] [3] | Group |
| Conditional Statistical Parity [10] | Group |
| Conditional Use Accuracy Equality* [12] | Group |
| Treatment Equality* [12] | Group |
| Test Fairness [18] [19] | Group |
| Construct Space [20] | Group |
| Statistical Parity [9] | Group |
| Minimax Pareto Fairness [21] | Group |
| Mulicalibration [22] | Subgroup |
| Subgroup Fairness [23] | Subgroup |
| Intersectional/Differential Fairness [24, 25] | Subgroup |
| Fairness Through Awareness [26] | Individual |
| Metric Fairness/Individual Fairness [27] | Individual |
| Fairness Elicitation [28] | Individual |
| Counterfactual Fairness [29] | Individual |
| Context-aware Fairness [30], [31] | n/a |
| Positionality-aware Fairness [32] | n/a |

A list of prominent fairness definitions used in algorithmic fairness literature, classified by their respective grouping types (e.g., group, subgroup, or individual fairness). Each definition is listed alongside alternative terminology from the literature and key citations that established or popularised the concept. The grouping type indicates whether the fairness definition is applied at the group, subgroup, or individual level. Definitions marked with an asterisk (*) are those selected for the current analysis.

Several fairness metrics were excluded. For instance, statistical parity and demographic parity were not used due to their assumption of equal base rates across groups—an assumption not valid for CVD, which occurs more frequently in males. Instead, definitions prioritising performance differences across groups were selected.

The selection focused on two core principles:

1. **Focus on group fairness:** Clinical decision-making often depends on demographic characteristics like age and sex. Thus, group-level fairness definitions are more interpretable and actionable than individual-level

metrics. This aligns with literature recommending group-level metrics in clinical risk tools.[3, 17]

2. **Importance of false negative rates:** In preventative medicine, avoiding false negatives is essential to minimise missed cases of high-risk individuals. Metrics such as equal opportunity and treatment equality directly address disparities in false negative rates/false negatives, aligning with clinical objectives.[33, 34]

From the initial shortlist, four definitions were retained to ensure interpretability and reduce redundancy in results: equal accuracy, equal opportunity, conditional use accuracy equality, and treatment equality. Although common in risk-prediction studies excluded calibration-based fairness definitions as they are only applicable to continuous risk predictions models, the thresholded individual risk factor distributions and PRS-augmented high-risk definitions are both binary predictions making calibration unsuitable. To evaluate disparities in these fairness definitions, we quantify fairness in each criteria using the using the following metrics (Table S2): accuracy equality difference (AED), equal opportunity difference (EOD), conditional use accuracy equality difference (CUAED), and treatment equality difference (TED).

Table S2: **Fairness quantifications for prediction models.**

| Fairness quantification | Quantification description | Quantification equation |
|---|---|---|
| Accuracy equality difference (AED) | Greatest difference in accuracy between any two groups within a characteristic | $\max_{i \neq j \in \{A,B,\ldots,N\}} \lvert Acc_i - Acc_j \rvert$ |
| Equal opportunity difference (EOD) | Greatest difference between the FNRs of any two groups within a characteristic | $\max_{i \neq j \in \{A,B,\ldots,N\}} \lvert FNR_i - FNR_j \rvert$ |
| Conditional use accuracy equality difference (CUAED) | Greatest difference between PPV or NPV of any two groups, whichever difference is greater, within a characteristic | $\max(\max_{i \neq j \in \{A,B,\ldots,N\}} \lvert PPV_i - PPV_j \rvert, \max_{i \neq j \in \{A,B,\ldots,N\}} \lvert NPV_i - NPV_j \rvert)$ |
| Treatment equality difference (TED) | Greatest difference between the FN:FP ratio of any two groups within a characteristic | $\max_{i \neq j \in \{A,B,\ldots,N\}} \lvert FN_i : FP_i - FN_j : FP_j \rvert$ |

Each quantification describes a method to measure fairness disparities across demographic groups $i, j \in \{A, B, \ldots, N\}$. The quantifications include differences in AED, EOD, CUAED, TED, calibration equality difference, and calibration slope equality difference. These metrics capture the maximum disparities in performance metrics across groups within each characteristic, providing a quantitative basis for assessing and comparing model fairness.

##### A.1.2 Fairness Assessment of Individual risk-factors: matching procedure and metrics

To minimise confounding by intersectional differences in demographics, a one-to-one nearest-neighbour (NN) matching procedure was applied separately for each group of interest within a characteristic $C$. For a given in-group $G_{\text{in}}$ (all individuals with a specific subgroup of $C$), matching was performed against the complementary out-group $G_{\text{out}} = X \setminus G_{\text{in}}$. Matching used only non-focal covariates (i.e., all variables except $C$) and was conducted *without replacement* so that each out-group individual was used at most once within a run.

Categorical covariates were one-hot encoded, and continuous covariates were standardised to zero mean and unit variance prior to matching. Euclidean distance on this preprocessed covariate set (excluding $C$) was used to quantify similarity. For individuals $x_i \in G_{\text{in}}$ and $x_j \in G_{\text{out}}$ with covariate vectors $\tilde{\boldsymbol{x}}_i$ and $\tilde{\boldsymbol{x}}_j$ (non-focal features only), the distance was

$$d(x_i, x_j) = \left\lVert \tilde{\boldsymbol{x}}_i - \tilde{\boldsymbol{x}}_j \right\rVert_2 = \sqrt{\sum_{k \neq C} \left(\tilde{x}_{ik} - \tilde{x}_{jk}\right)^2}.$$

**Algorithm 1** One-to-one nearest neighbours matching

**Require:** Population $X$; characteristic $C$; in-group $G_{\text{in}} \subseteq X$; out-group $G_{\text{out}} = X \setminus G_{\text{in}}$; preprocessed non-focal features $\tilde{\boldsymbol{x}}$

**if** $|G_{\text{in}}| > |G_{\text{out}}|$ **then**<br>
  Subsample $G_{\text{in}}$ to size $|G_{\text{out}}|$ (fixed seed)<br>
**end if**<br>
Fit $k$NN index on $G_{\text{out}}$ using $\tilde{\boldsymbol{x}}$; mark all $G_{\text{out}}$ as unused<br>
Partition $G_{\text{in}}$ into chunks; initialise matching $M \leftarrow \emptyset$<br>
**for** each chunk $\mathcal{S} \subseteq G_{\text{in}}$ **do**<br>
  $R \leftarrow \mathcal{S}$;  $k \leftarrow k_0$<br>
  **while** $R \neq \emptyset$ **do**<br>
    For each $x_i \in R$, retrieve its $k$ nearest neighbours in $G_{\text{out}}$<br>
    Greedily assign the nearest *unused* neighbour to each $x_i$; mark assigned controls as used<br>
    $R \leftarrow$ set of $x_i$ still unmatched<br>
    **if** $R = \emptyset$ **then break**<br>
    **else**$k < |G_{\text{out}}|$<br>
      $k \leftarrow \min(2k,\ |G_{\text{out}}|)$<br>
    **end if**<br>
  **end while**<br>
**end for**<br>
**return** $M$

For each group $g$ in a characteristic $C$, define the in-group $G_{\text{in}} = \{x \in X : C(x) = g\}$ and the out-group $G_{\text{out}} = X \setminus G_{\text{in}}$. Matching uses only non-focal covariates (preprocessed upstream), denoted $\boldsymbol{z}(x)$. If $|G_{\text{in}}| > |G_{\text{out}}|$, $G_{\text{in}}$ is randomly downsampled (fixed seed) to $|G_{\text{out}}|$ to permit one-to-one matching. A $k$-nearest-neighbour index (Euclidean distance) is fit on $G_{\text{out}}$ using $\boldsymbol{z}(x)$. In-group individuals are processed in chunks; within each chunk, a greedy assignment selects for each $x_i \in G_{\text{in}}$ the nearest *unused* neighbour in $G_{\text{out}}$, starting with $k = k_0$ and doubling $k$ until $k = |G_{\text{out}}|$. When $k$ spans the full pool, the remaining unmatched $x_i$ are completed by selecting their nearest unused neighbours from the remaining controls, yielding a one-to-one, without-replacement matching for the (possibly downsampled) $G_{\text{in}}$. The procedure is repeated across randomly-ordered bootstrap samples (to prevent earlier-indexed samples having better neighbour matches), and fairness metrics are computed on the matched pairs for each characteristic.

Table S3: **Fairness quantifications for individual risk factors.**

| Fairness quantification | Quantification description | Quantification equation |
|---|---|---|
| Accuracy equality difference (AED) | Greatest difference in accuracy between any matched in- and out-group within a characteristic | $\max_{(G_{in},G_{out})\in\{A,B,\ldots,N\}} \lvert Acc(G_{in}) - Acc(G_{out})\rvert$ |
| Equal opportunity difference (EOD) | Greatest difference between the FNRs of any matched in- and out-group within a characteristic | $\max_{(G_{in},G_{out})\in\{A,B,\ldots,N\}} \lvert FNR(G_{in}) - FNR(G_{out}))\rvert$ |
| Conditional use accuracy equality difference (CUAED) | Greatest difference between PPV or NPV of any matched in- and out-group, whichever difference is greater, within a characteristic | $\max(\max_{(G_{in},G_{out})\in\{A,B,\ldots,X\}} \lvert PPV(G_{in}) - PPV(G_{out})\rvert,$ $\max_{(G_{in},G_{out})\in\{A,B,\ldots,N\}} \lvert NPV(G_{in}) - NPV(G_{out})$ |
| Treatment equality difference (TED) | Greatest difference between the FN:FP ratio of any matched in- and out-group within a characteristic | $\max_{(G_{in},G_{out})\in\{A,B,\ldots,N\}} \lvert FN(G_{in}) : FP(G_{in}) - FN(G_{out}) : FP(G_{out})\rvert$ |

Fairness quantifications for individual risk factors, calculated within each characteristic C (accuracy equality difference (AED), equal opportunity difference (EOD), conditional use accuracy equality difference (CUAED), and treatment equality difference (TED)). Each quantification measures the greatest difference between any matched in-group and out-group within the characteristic. Equations are defined for each quantification metric (larger values indicate greater disparities).

#### A.2 Supplementary Figures

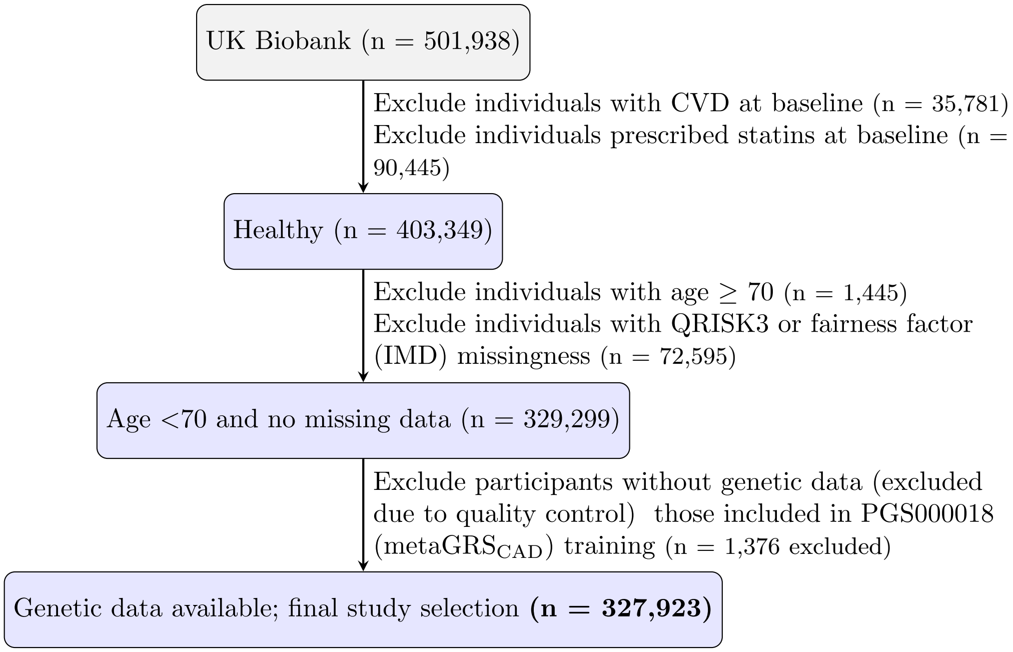


Figure S1: **Flowchart of study inclusion criteria for single risk factor fairness analysis.** The stepwise process for participant selection from the UK Biobank cohort (n=501,938) is outlined. Individuals with CVD or prescribed statins at baseline, those with missing key predictor data (risk factors for QRISK3 and IMD), or with age >70 were excluded. Additionally, those with missing genetic data due to quality control were excluded (n=13,089). This resulted in a final study sample (n = 327,923) for subsequent analysis.

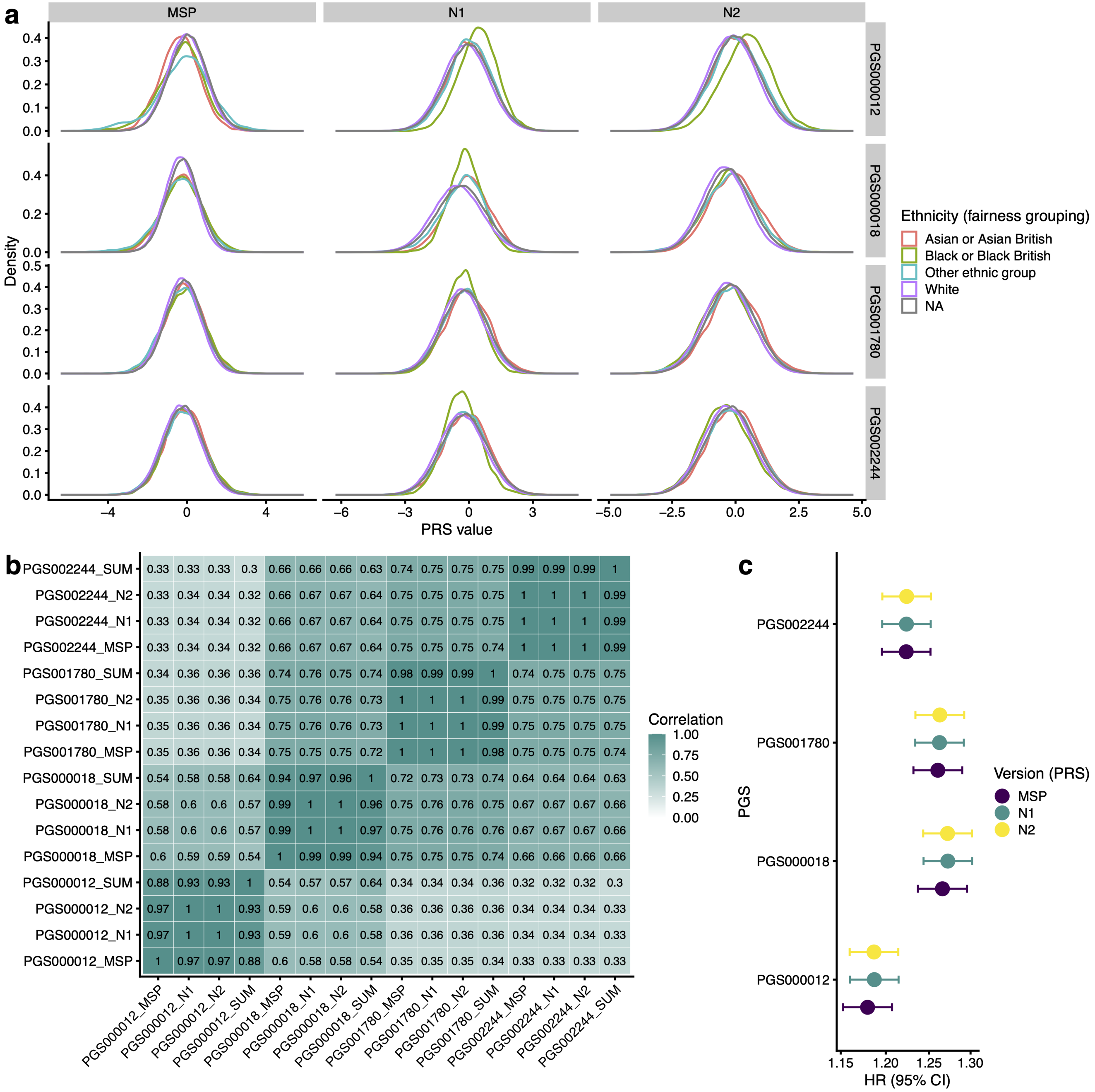


Figure S2: **Summary of the evaluated PRS and normalisation methods. (a) Density plots of PRS distributions for the four evaluated scores and three evaluated normalisation methods (MSP, N1, N2), stratified by the Ethnicity groupings used in fairness analysis.** (b) Spearman correlation between evaluated PRS and normalisation methods for all individuals with genetic data. (c) Association of each PGS and normalisation version with incident CVD events (within 10-years). Association is measured as an Hazard Ratio (OR) per SD change the PGS (and 95% confidence interval) using sex-stratified Cox-proportional hazards regression, adjusted for age at baseline.

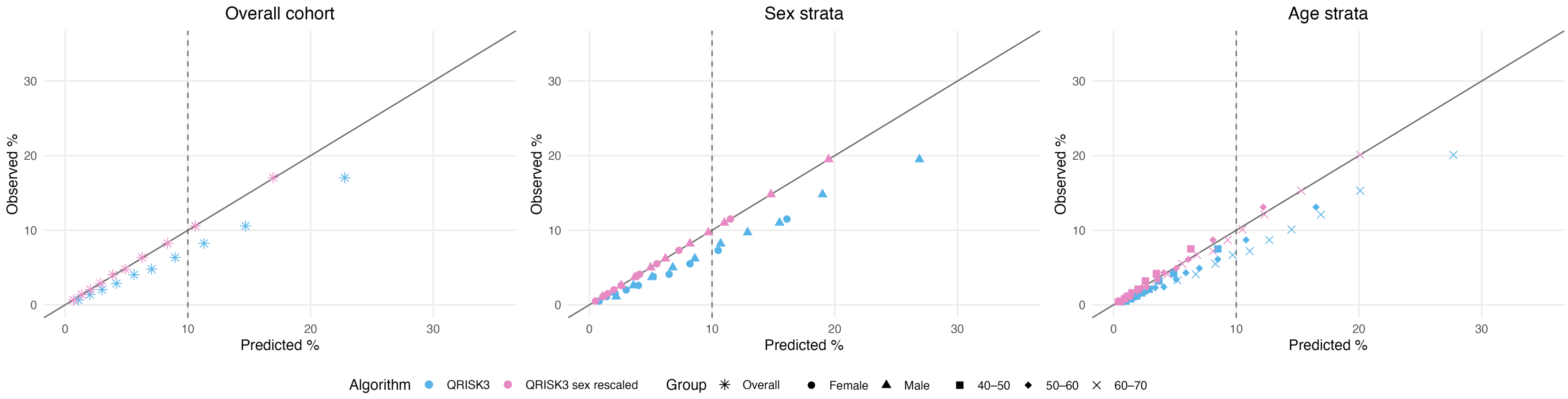


Figure S3: **Recalibration of QRISK3 predictions by sex.** a) Observed versus predicted 10-year cardiovascular disease (CVD) risk for the original QRISK3 model (blue) and after sex-specific rescaling (pink). The diagonal line represents perfect calibration. The vertical dashed line indicates the decision risk threshold (10%). Points represent deciles of predicted risk, with corresponding observed event rates. b) Calibration of QRISK3 before and after sex-specific rescaling across age deciles. Points represent mean predicted 10-year CVD risk versus observed risk (from Kaplan–Meier estimates) within deciles of age in the study population.

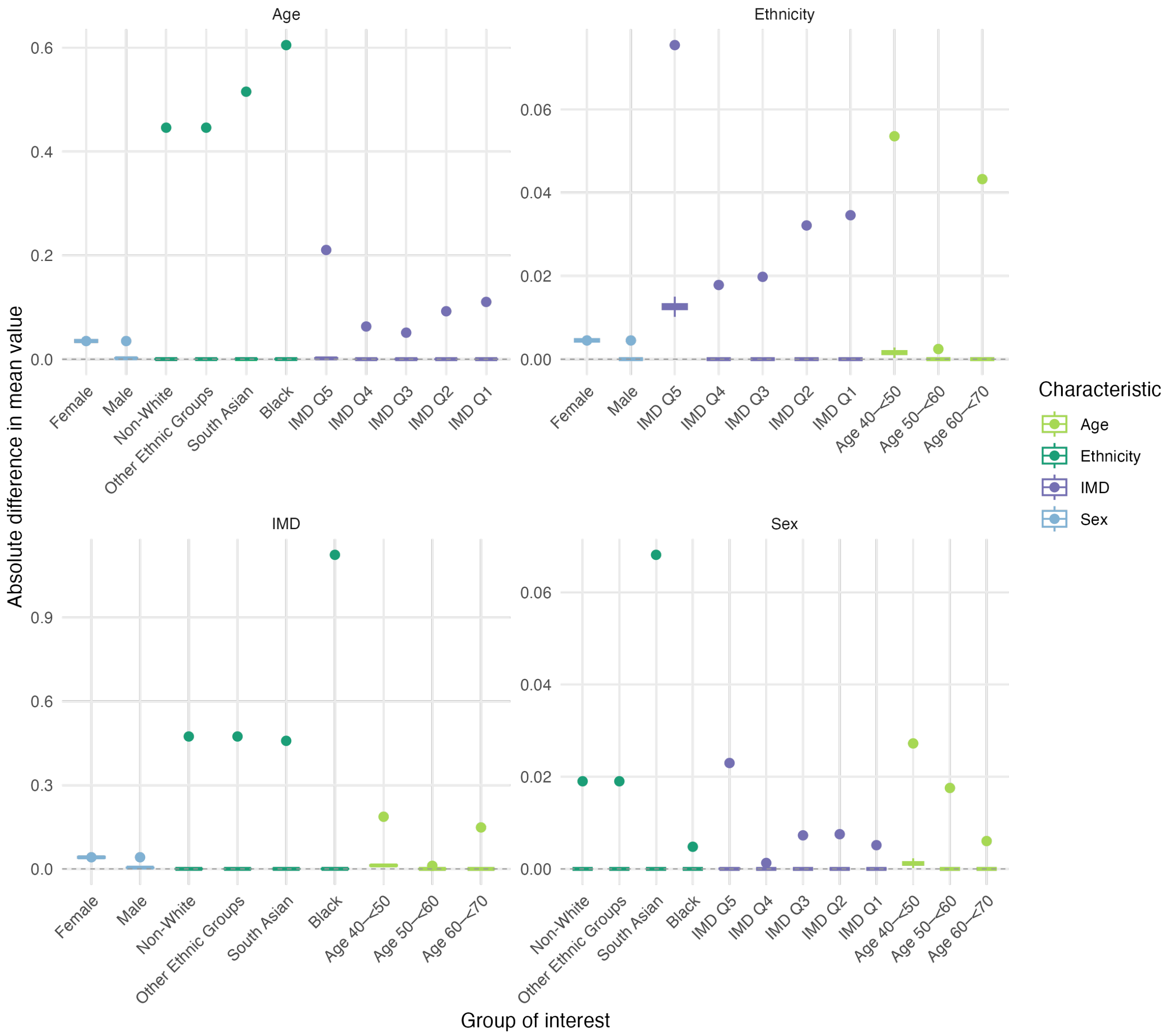


Figure S4: **Differences between the mean value in the group and its matched group, and the group and its unmatched out-group in the UKB data, across all characteristics.** Effectiveness of matching procedure is shown with a comparison for each group of: 1) the difference between the means of each in-group and its 1:1 matched out-group (boxplots), with 2) the difference between means of each in-group and the unmatched out-group in the UK Biobank data (scatter points). A one-hot-encoding and distance metric was defined to measure the distance between ethnicity and sex groups, since these are categorical values. The colour denotes the characteristic to which each group of interest belongs. Larger values indicate more difference between groups; boxplots with values close to 0 indicate perfect matching across bootstraps.

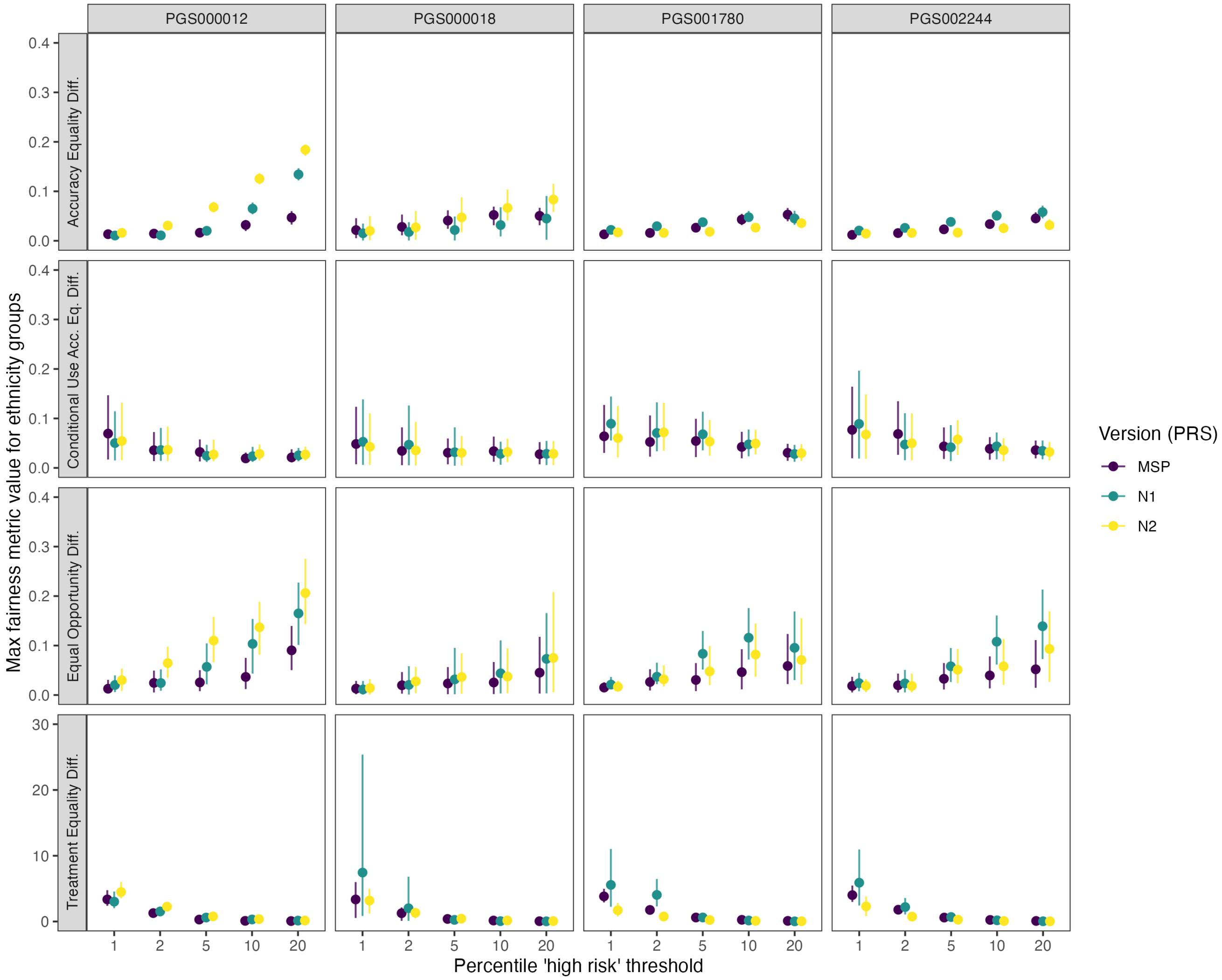


Figure S5: **Fairness metrics for ethnicity-matched groups across PRSs and normalisation versions.** Shown are the maximum differences in four fairness metrics (accuracy equality, conditional use accuracy equality, equal opportunity, treatment equality) by PRS (columns) and high-risk threshold (x-axis: top 1–20%). Points show bootstrap means with 95% confidence intervals; colours indicate normalisation versions (MSP, N1, N2).

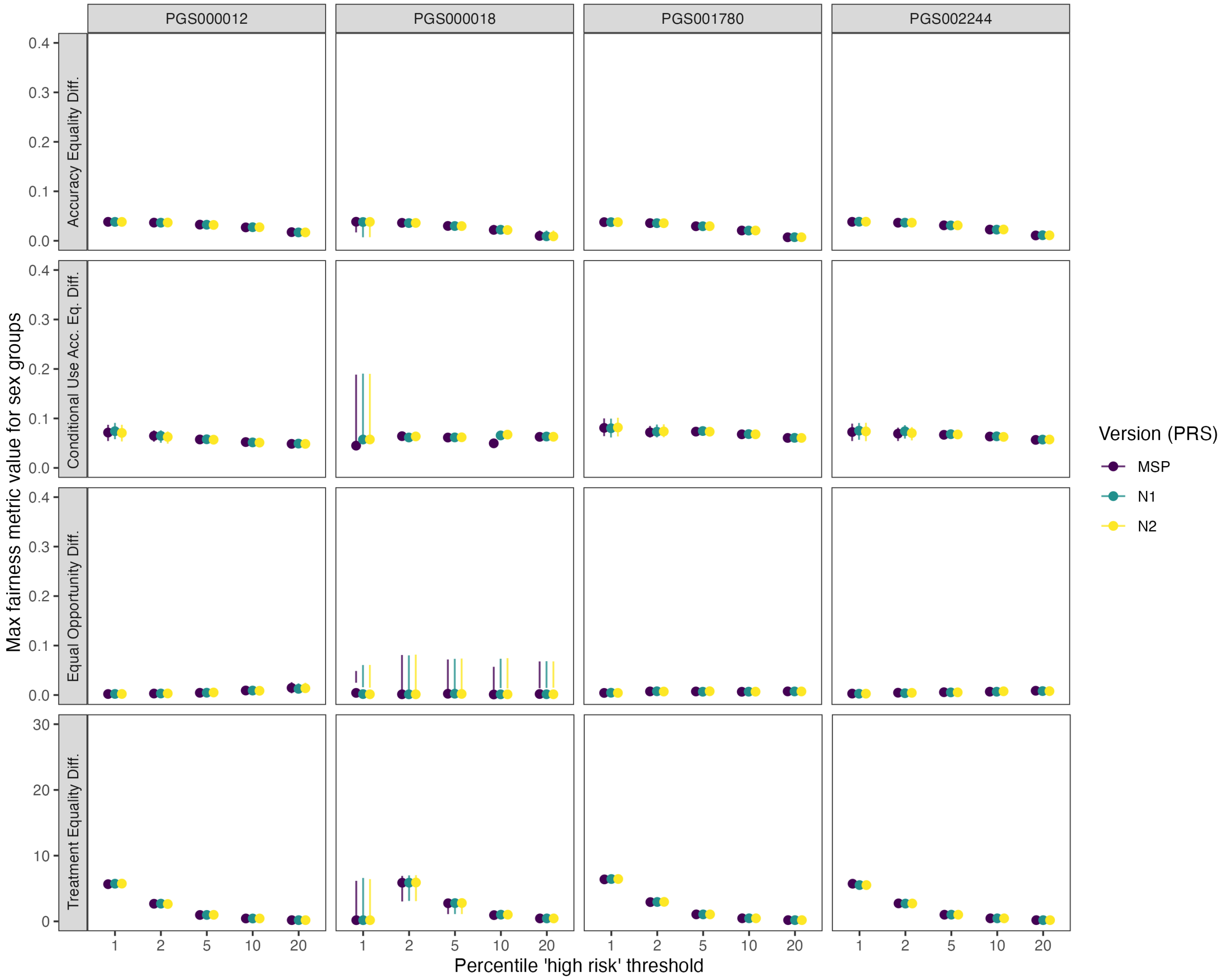


Figure S6: **Fairness metrics for sex-matched groups across PRSs and normalisation versions.** Shown are the maximum differences in four fairness metrics (accuracy equality, conditional use accuracy equality, equal opportunity, treatment equality) by PRS (columns) and high-risk threshold (x-axis: top 1–20%). Points show bootstrap means with 95% confidence intervals; colours indicate normalisation versions (MSP, N1, N2).

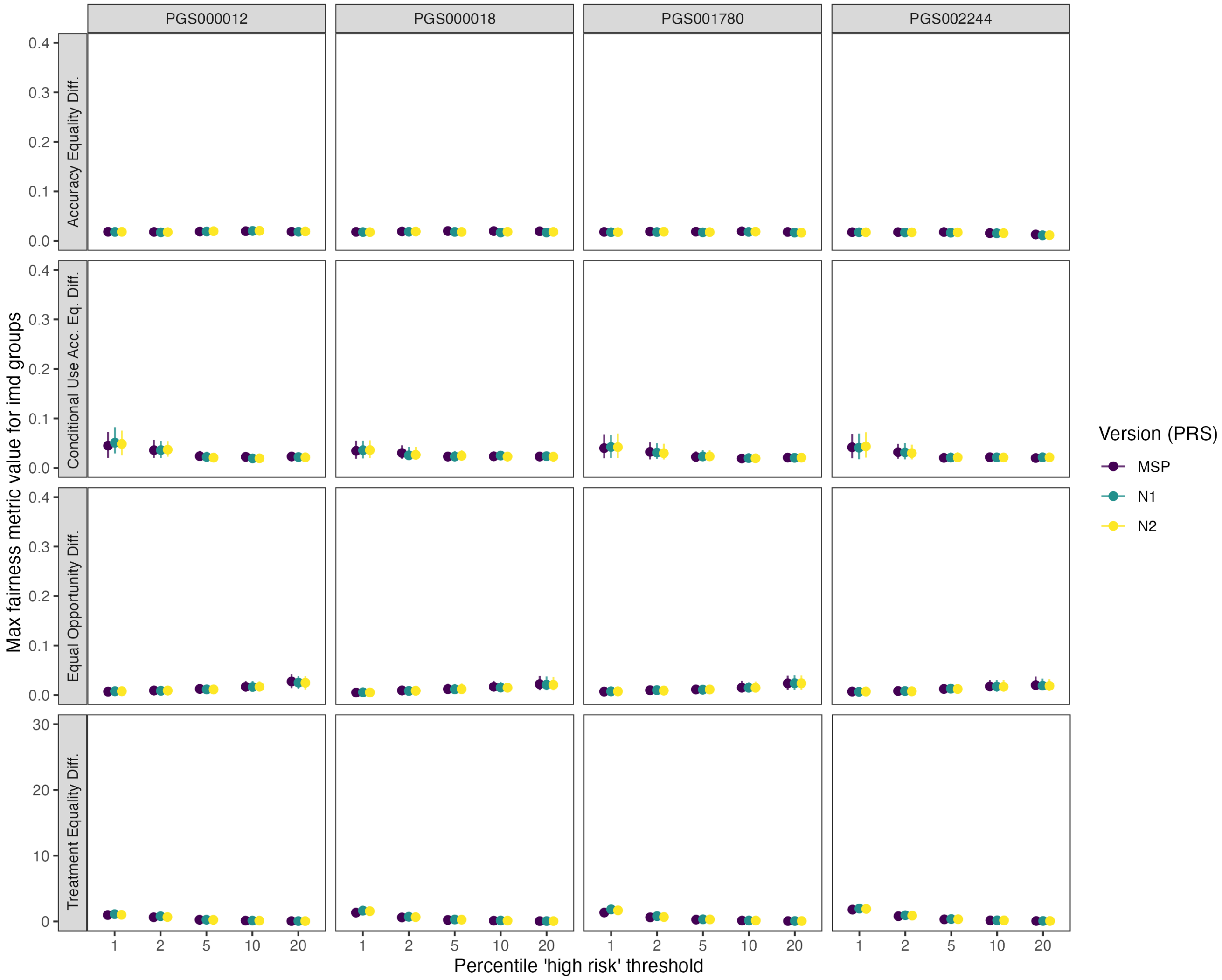


Figure S7: **Fairness metrics for imd-matched groups across PRSs and normalisation versions.** Shown are the maximum differences in four fairness metrics (accuracy equality, conditional use accuracy equality, equal opportunity, treatment equality) by PRS (columns) and high-risk threshold (x-axis: top 1–20%). Points show bootstrap means with 95% confidence intervals; colours indicate normalisation versions (MSP, N1, N2).

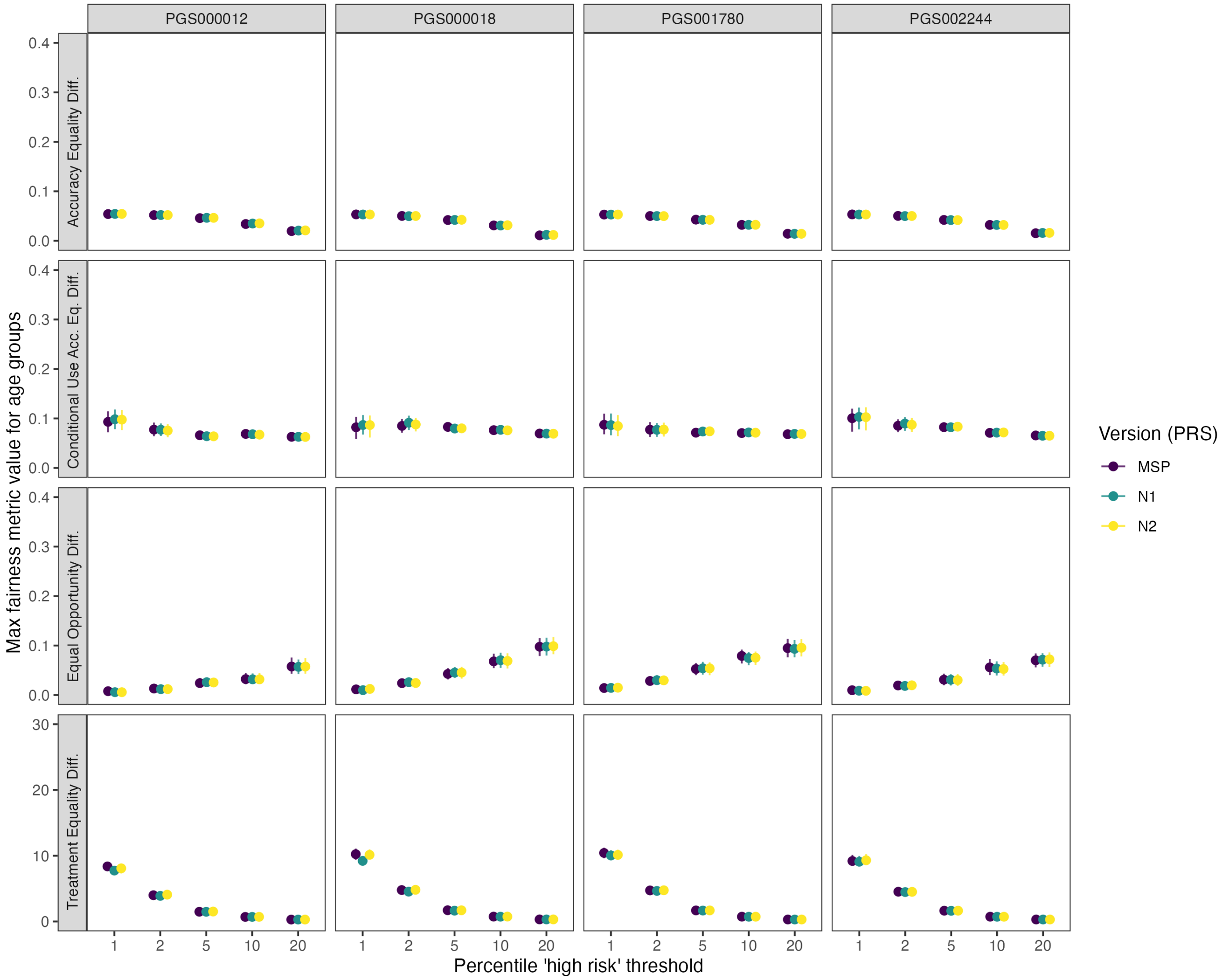


Figure S8: **Fairness metrics for age-matched groups across PRSs and normalisation versions.** Shown are the maximum differences in four fairness metrics (accuracy equality, conditional use accuracy equality, equal opportunity, treatment equality) by PRS (columns) and high-risk threshold (x-axis: top 1–20%). Points show bootstrap means with 95% confidence intervals; colours indicate normalisation versions (MSP, N1, N2).

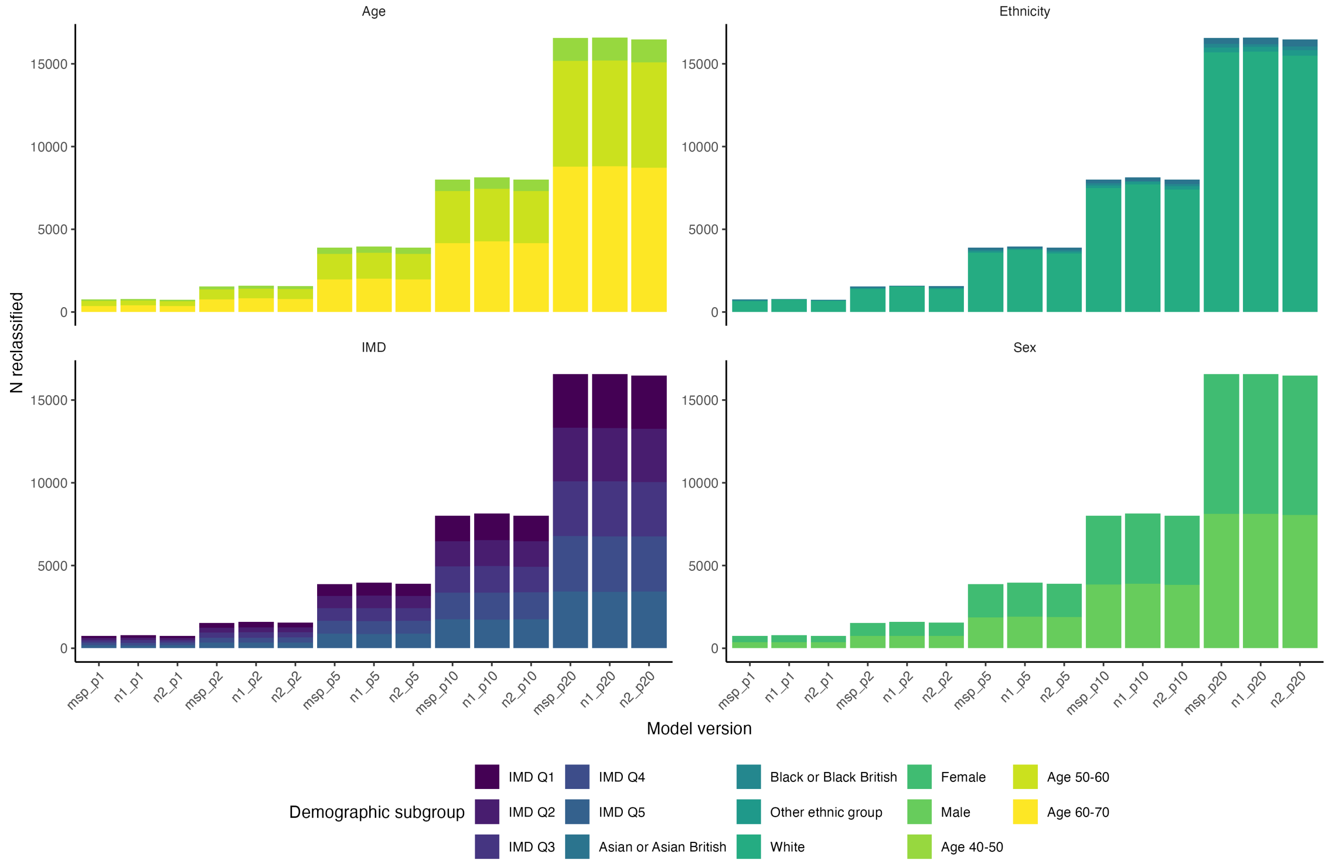


Figure S9: **Demographic groups of the individuals reclassified into CVD high risk group with the addition of PRS (PGS000018) to the baseline QRISK3 models.** The x-axis shows the specific model: PRS normalisation (N1, N2, MSP); baseline QRISK3 version (sex-recalibrated, sex-ethnicity recalibrated indicated with “sex” and “eth” respectively); and PRS percentile (1, 2, 5, 10, 20) used as the threshold to determine reclassification into the high risk category. The y axis shows the count of reclassified individuals in each demographic group. The demographic group membership is indicated by the colours. There is one subplot for each characteristic (age, ethnicity, IMD, sex).

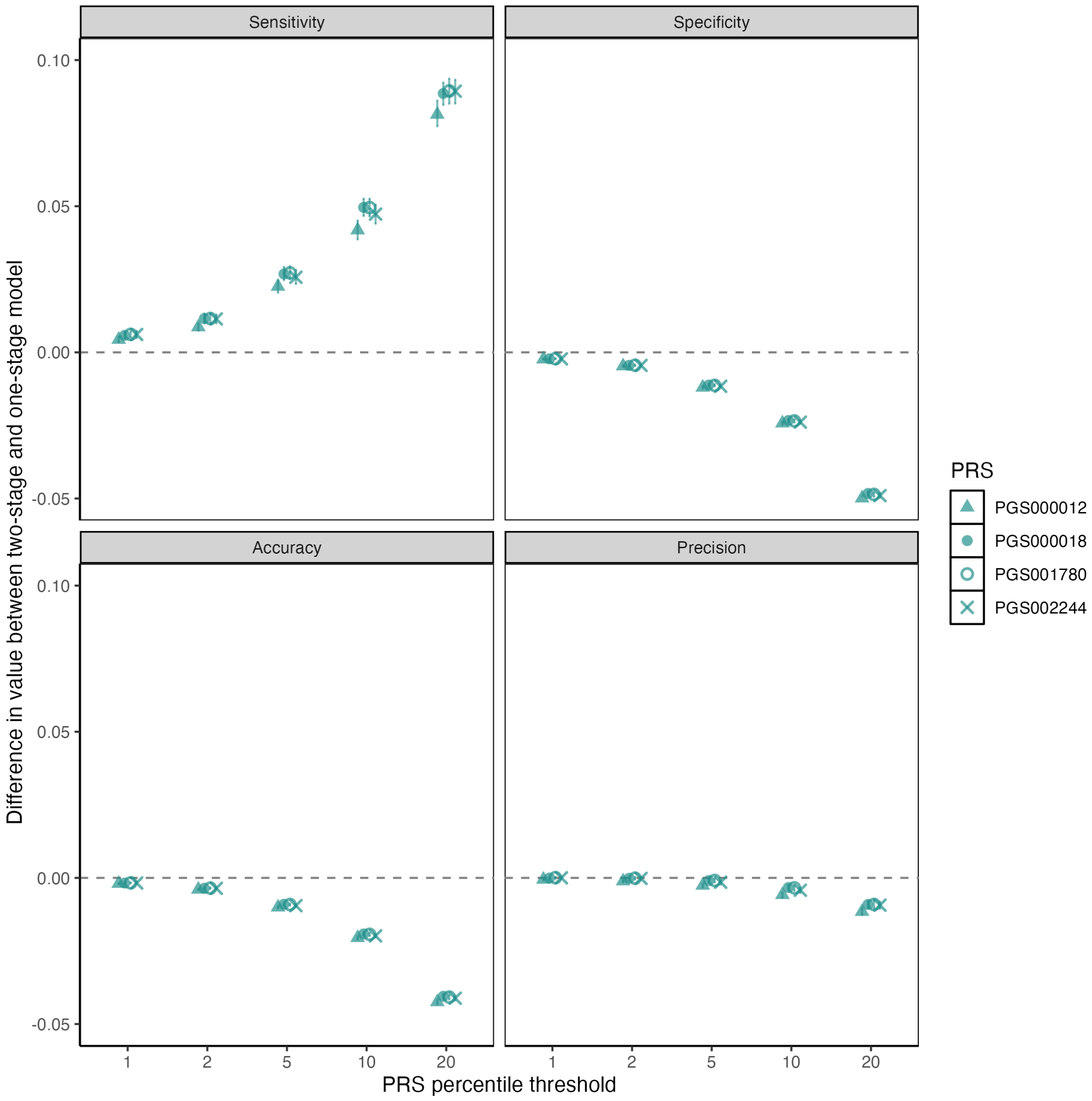


Figure S10: **Difference in performance between two-stage PRS-added model and one-stage model for PRS percentile thresholds for all PRS at normalisation N1.** Sensitivity, specificity, accuracy, and precision between two-stage and one-stage models are presented alongside 95% confidence intervals calculated across 200 bootstraps. Changes in overall performance were evaluated for the QRISK3 sex-recalibrated baseline model and their two-stage PRS-added versions. This was done for all PRSs (PGS000012, PGS000018, PGS001780, PGS002244) normalisation N1 across all PRS percentile thresholds. Results are mostly consistent across PRS, but sensitivity differs, mimicking the differences in effect size of the scores (Figure S2).

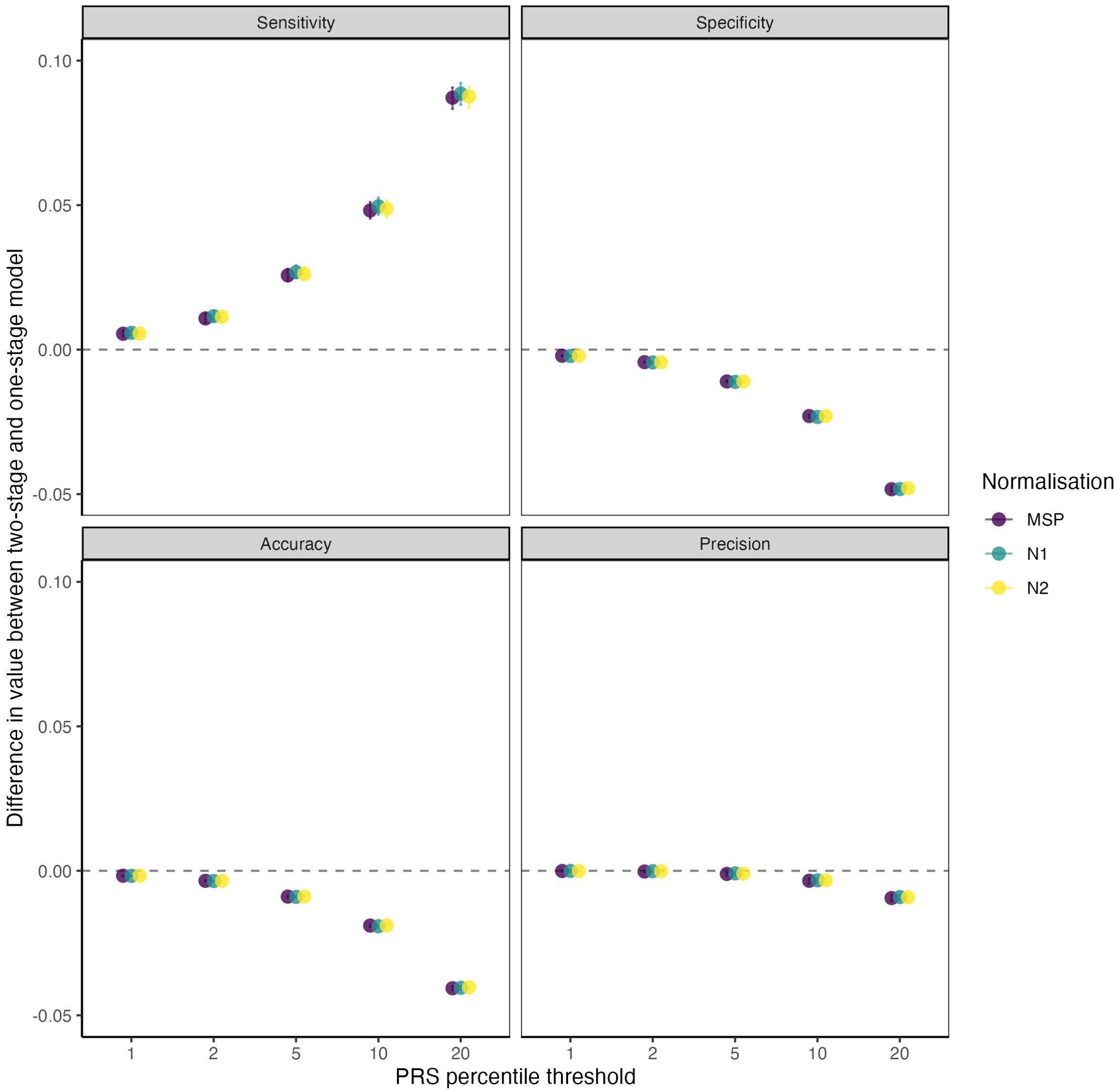


Figure S11: **Difference in performance between two-stage PRS-added model and one-stage model for PRS percentile thresholds for PGS000018 for all normalisations.** Sensitivity, specificity, accuracy, and precision between two-stage and one-stage models are presented alongside 95% confidence intervals calculated across 200 bootstraps. Changes in overall performance were evaluated for the QRISK3 sex-recalibrated baseline model and their two-stage PRS-added versions. This was done for PGS000018 for all normalisations (MSP, N1, N2) across al percentile thresholds. Results are consistent across normalisations.

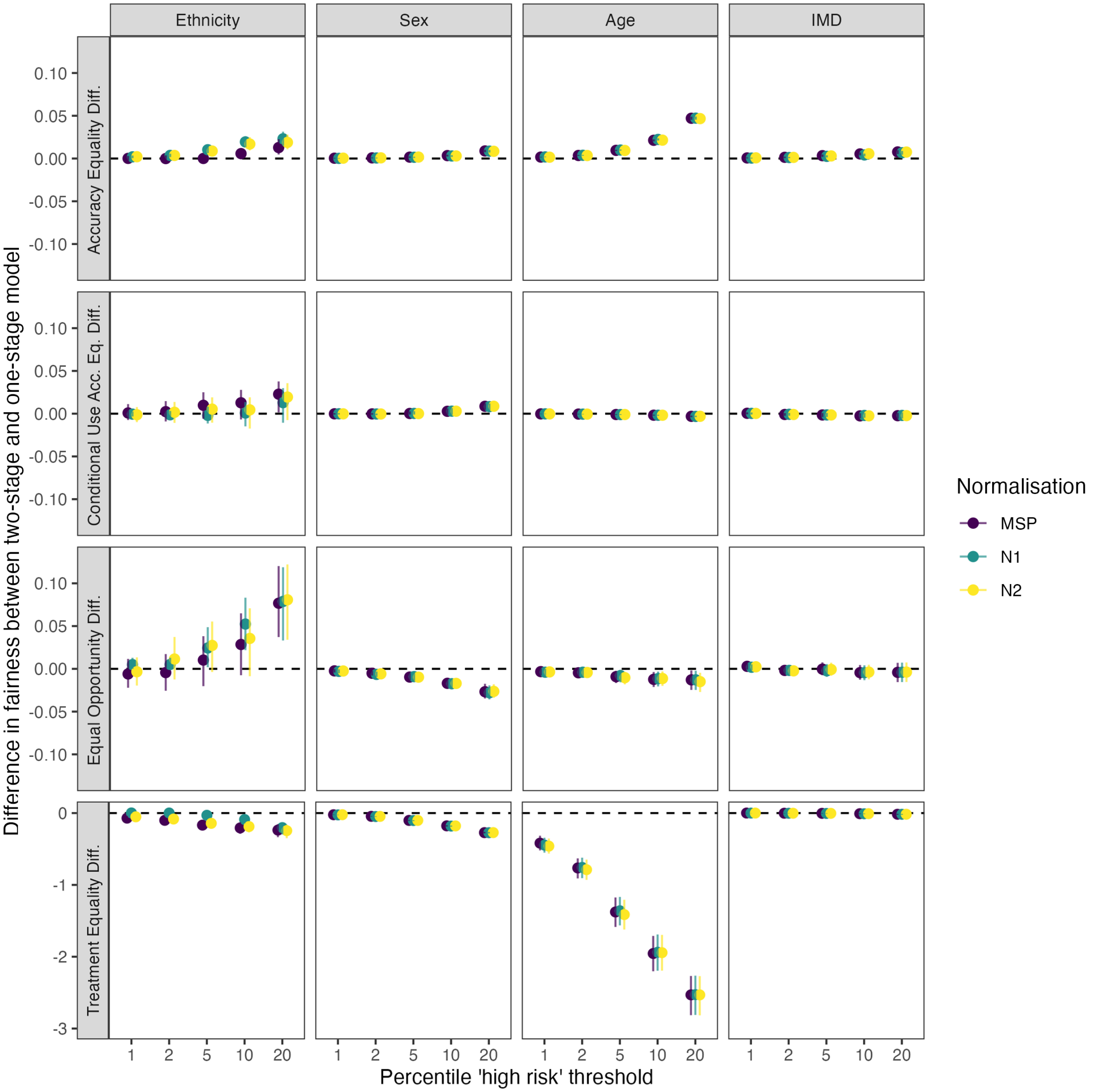


Figure S12: **Difference in fairness metric values between two-stage PRS-added model and one-stage model for PRS percentile thresholds for all normalisations of PGS000018.** Mean difference in values for AED, CUAED, EOD, and TED between two-stage and one-stage models are presented alongside 95% confidence intervals calculated across 200 bootstraps. Changes in metrics for ethnicity, sex, age, and IMD characteristics were evaluated for the QRISK3 sex-recalibrated baseline model and the two-stage PRS-added version. This was done across PRS percentile thresholds for PGS000018 for 3 normalisation versions: MSP, N1, and N2. Results were mostly consistent across normalisations, with some small differences for ethnicity in EOD and TED.

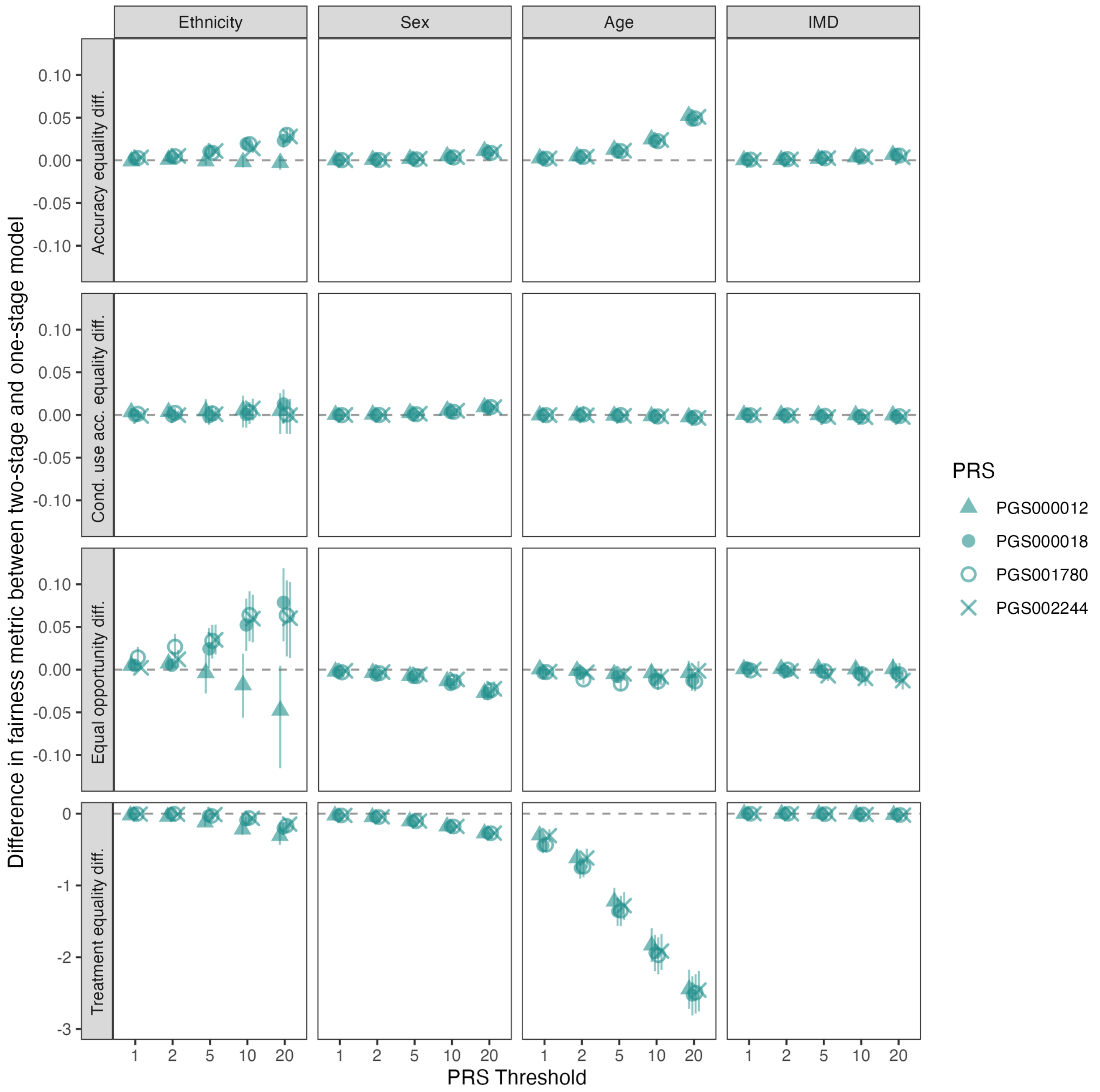


Figure S13: **Difference in fairness metric values between two-stage PRS-added model and one-stage model for PRS percentile thresholds for all PRS versions.** Mean difference in values for AED, CUAED, EOD, and TED between two-stage and one-stage models are presented alongside 95% confidence intervals calculated across 200 bootstraps. Changes in metrics for ethnicity, sex, age, and IMD characteristics were evaluated for the QRISK3 sex-recalibrated baseline model and the two-stage PRS-added version. This was done across PRS percentile thresholds for 4 PRS versions: PGS000012, PGS000018, PGS001780, PGS002244. Results were mostly consistent across PRSs, with some differences observed for PGS000012 for TED for ethnicity groups, especially at higher thresholds.

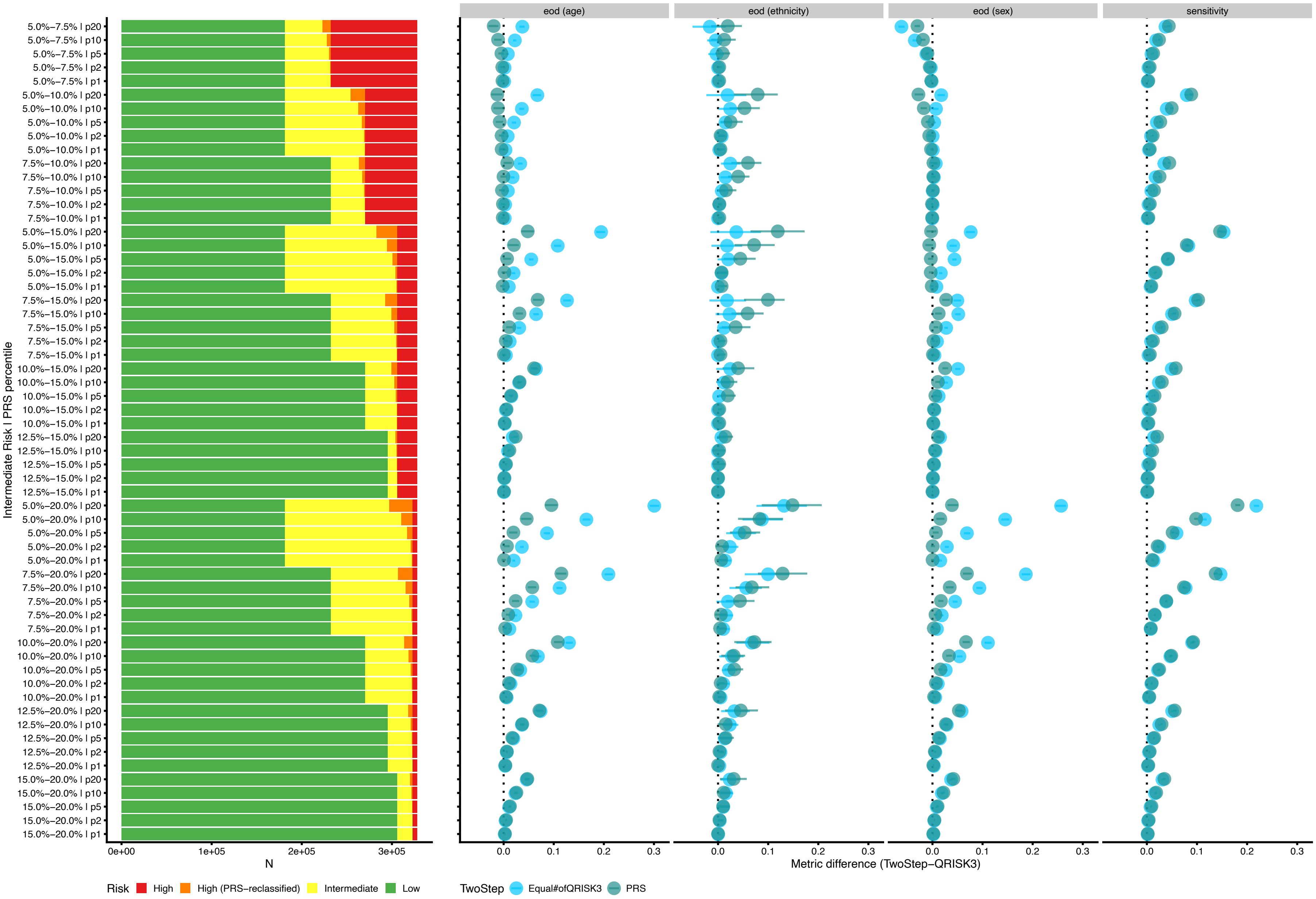


Figure S14: **Sensitivity analysis varying risk threshold (part 1).** Absolute numbers of individuals reclassified by PRS percentile groups together with accuracy equality difference (AED) and sensitivity differences between the TwoStep PRS-reclassification and Equal number of QRISK3 models across age, sex, and ethnicity strata, evaluated under alternative medium-risk thresholds.

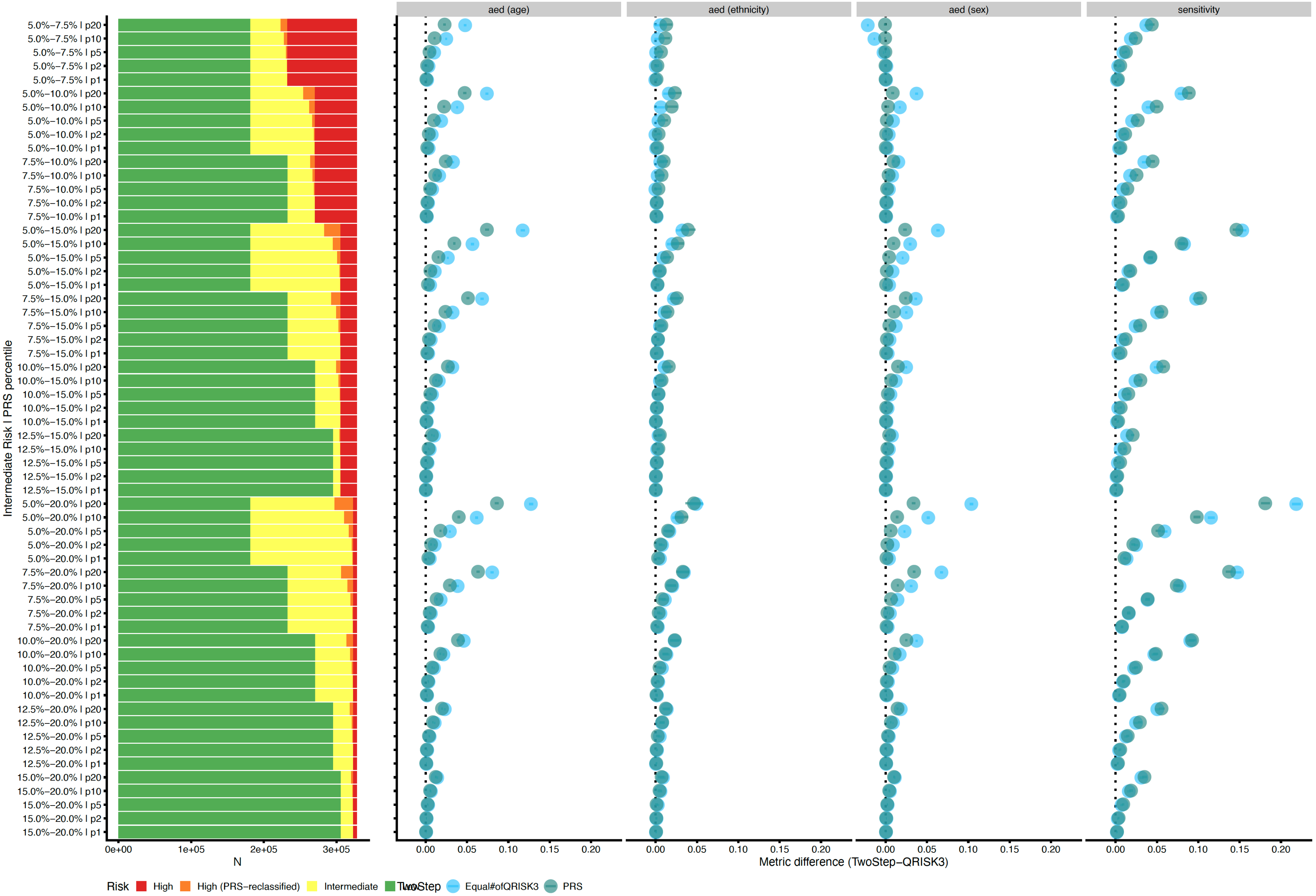


Figure S15: **Sensitivity analysis varying risk threshold (part 2).** Absolute numbers of individuals reclassified by PRS percentile groups together with equal opportunity difference (EOD) and sensitivity differences between the TwoStep PRS-reclassification and Equal number of QRISK3 models across age, sex, and ethnicity strata, evaluated under alternative medium-risk thresholds.

#### A.3 Supplementary Tables

Table S4: **Codes used to define incident and prevalent CVD in UK Biobank.**

| Source | Code | Description |
|---|---|---|
| HES (ICD-10) | G45 | Transient cerebral ischaemic attacks and related syndromes |
| HES (ICD-10) | I20 | Angina pectoris |
| HES (ICD-10) | I21 | Acute myocardial infarction |
| HES (ICD-10) | I22 | Subsequent myocardial infarction |
| HES (ICD-10) | I23 | Certain current complications following acute myocardial infarction |
| HES (ICD-10) | I24 | Other acute ischaemic heart diseases |
| HES (ICD-10) | I25 | Chronic ischaemic heart disease |
| HES (ICD-10) | I25.2 | Old myocardial infarction |
| HES (ICD-10) | I63 | Cerebral infarction |
| HES (ICD-10) | I64 | Stroke, not specified as haemorrhage or infarction |
| HES (ICD-9) | 410 | Acute myocardial infarction |
| HES (ICD-9) | 411 | Other acute and subacute forms of ischemic heart disease |
| HES (ICD-9) | 412 | Old myocardial infarction |
| HES (ICD-9) | 413 | Angina pectoris |
| HES (ICD-9) | 414 | Other forms of chronic ischemic heart disease |
| HES (ICD-9) | 434 | Occlusion of cerebral arteries |
| HES (ICD-9) | 436 | Acute but ill-defined cerebrovascular disease |
| HES procedure (OPCS-4) | K40.2 | Saphenous vein graft replacement of 2 coronary arteries |
| HES procedure codes (OPCS-4) | K40.1 | Saphenous vein graft replacement of 1 coronary artery |
| HES procedure codes (OPCS-4) | K40.3 | Saphenous vein graft replacement of 3 coronary arteries |
| HES procedure codes (OPCS-4) | K40.4 | Saphenous vein graft replacement of 4 or more coronary arteries |
| HES procedure codes (OPCS-4) | K40.8 | Saphenous vein graft replacement of coronary artery: Other |
| HES procedure codes (OPCS-4) | K40.9 | Saphenous vein graft replacement of coronary artery: Unspecified |
| HES procedure codes (OPCS-4) | K41.1 | Autograft replacement of 1 coronary artery NEC |
| HES procedure codes (OPCS-4) | K41.2 | Autograft replacement of 2 coronary arteries NEC |
| HES procedure codes (OPCS-4) | K41.3 | Autograft replacement of 3 coronary arteries NEC |
| HES procedure codes (OPCS-4) | K41.4 | Autograft replacement of 4 or more coronary arteries NEC |
| HES procedure codes (OPCS-4) | K41.8 | Other autograft replacement of coronary artery: Other |
| HES procedure codes (OPCS-4) | K41.9 | Other autograft replacement of coronary artery: Unspecified |
| HES procedure codes (OPCS-4) | K42 | Allograft replacement of coronary artery |
| HES procedure codes (OPCS-4) | K43 | Prosthetic replacement of coronary artery |
| HES procedure codes (OPCS-4) | K44 | Other replacement of coronary artery |
| HES procedure codes (OPCS-4) | K45.1 | Double anastomosis of mammary arteries to coronary arteries |
| HES procedure codes (OPCS-4) | K45.2 | Double anastomosis of thoracic arteries to coronary arteries NEC |
| HES procedure codes (OPCS-4) | K45.3 | Anastomosis of mammary artery to left anterior descending coronary artery |
| HES procedure codes (OPCS-4) | K45.4 | Anastomosis of mammary artery to coronary artery NEC |
| HES procedure codes (OPCS-4) | K45.5 | Anastomosis of thoracic artery to coronary artery NEC |
| HES procedure codes (OPCS-4) | K45.6 | Revision of connection of thoracic artery to coronary artery |
| HES procedure codes (OPCS-4) | K45.8 | Connection of thoracic artery to coronary artery: Other specified |
| HES procedure codes (OPCS-4) | K45.9 | Connection of thoracic artery to coronary artery: Unspecified |
| HES procedure codes (OPCS-4) | K46 | Other bypass of coronary artery |
| HES procedure codes (OPCS-4) | K47.1 | Endarterectomy of coronary artery |
| HES procedure codes (OPCS-4) | K49.1 | Percutaneous transluminal balloon angioplasty of 1 coronary artery |
| HES procedure codes (OPCS-4) | K49.2 | Percutaneous transluminal balloon angioplasty of multiple coronary arteries |
| HES procedure codes (OPCS-4) | K49.3 | Percutaneous transluminal balloon angioplasty of bypass graft of coronary artery |

| Continuation of Table S4 | | |
|---|---|---|
| Source | Code | Description |
| HES procedure codes (OPCS-4) | K49.4 | Percutaneous transluminal cutting balloon angioplasty of coronary artery |
| HES procedure codes (OPCS-4) | K49.8 | Transluminal balloon angioplasty of coronary artery: Other |
| HES procedure codes (OPCS-4) | K49.9 | Transluminal balloon angioplasty of coronary artery: Unspecified |
| HES procedure codes (OPCS-4) | K50 | Other therapeutic transluminal operations on coronary artery |
| HES procedure codes (OPCS-4) | K50.2 | Percutaneous transluminal coronary thrombolysis using streptokinase |
| HES procedure codes (OPCS-4) | K75.1 | Percutaneous transluminal balloon angioplasty and insertion of 1-2 drug-eluting stents into coronary artery |
| HES procedure codes (OPCS-4) | K75.2 | Percutaneous transluminal balloon angioplasty and insertion of 3 or more drug-eluting stents into coronary artery |
| HES procedure codes (OPCS-4) | K75.3 | Percutaneous transluminal balloon angioplasty and insertion of 1-2 stents into coronary artery |
| HES procedure codes (OPCS-4) | K75.4 | Percutaneous transluminal balloon angioplasty and insertion of 3 or more stents into coronary artery NEC |
| HES procedure codes (OPCS-4) | K75.8 | Percutaneous transluminal balloon angioplasty and insertion of stent into coronary artery: Other |
| HES procedure codes (OPCS-4) | K75.9 | Percutaneous transluminal balloon angioplasty and insertion of stent into coronary artery: Unspecified |
| UKB field | 2966 | Age high blood pressure diagnosed |
| UKB field | 3627 | Age angina diagnosed |
| UKB field | 3894 | Age heart attack diagnosed |
| UKB field | 4056 | Age stroke diagnosed |
| UKB field | 6150 | Vascular/heart problems doctor diagnosed: 1 heart attack, 3 stroke |
| UKB field | 6150 | Vascular/heart problems diagnosed by doctor: 2 angina |
| UKB field | 20002 | Self-reported: 1075 heart attack/myocardial infarction, 1082 transient ischaemic attack (TIA), 1583 ischaemic stroke |
| UKB field | 20004 | Operation code: 1070 coronary angioplasty (PTCA) +/- stent, 1095 coronary artery bypass grafts (CABG) |
| UKB field | 20004 | Operation code: 1071 other arterial surgery/revascularisation procedures, 1105 carotid artery surgery/endarterectomy, 1109 carotid artery angioplasty +/- stent, 1514 coronary angiogram |
| UKB field | 42006 | Date of stroke |
| UKB field | 42008 | Date of ischaemic stroke |

Table S6: **QRISK3 risk factors.**

| Risk factor | UKB field / source | Phenotyping details |
|---|---|---|
| Sex | UKB:31 | |
| Age | UKB:21022 | |
| Townsend deprivation index | UKB:22189 | |
| Height | UKB:50 | Used to calculate BMI |
| Weight | UKB:21002 | Used to calculate BMI |

| Continuation of Table S6 | | |
|---|---|---|
| Risk factor | UKB field / source | Phenotyping details |
| Ethnicity | UKB:21000 | Participants self-reporting their ethnicity as "White" (code 1), "British" (code 1001), "Irish" (code 1002), "Any other White background" (code 1003), "Prefer not to answer", had missing data in the ethnicity field were coded in the QRISK3 ethnicity category "White or not stated". Participants self-reporting "Indian" (code 3001), "Pakistani" (code 3002), "Bangladeshi" (code 3003), "Black Caribbean" (code 4001), "Black African" (code 4002), or "Chinese" (code 5) were kept as-is as these categories matched QRISK3 ethnicity categories. Participants self-reporting "Asian or Asian British" (code 3) or "Any other Asian background" (code 3004) were coded in the QRISK3 ethnicity category "Other Asian". All other self-reported ethnicities were coded in the QRISK3 category "Other ethnic group" |
| Smoking | UKB:3456 & UKB:20116 | Smoking status (non-smoker, ex-smoker, light smoker, moderate smoker, or heavy smoker) was defined based on both self-reported current smoking (field #20116) along with the number of cigarettes smoked per day for current smokers (field #3456). Participants with answering "don't know" or "prefer not to answer" to the smoking status field (field #20116) were treated as non-smokers. Current smokers who reported smoking less than 10 cigarettes per day were coded as light smokers. Current smokers who reported smoking between 10 and 20 cigarettes per day were coded as moderate smokers. Current smokers who reported smoking 20 or more cigarettes per day were coded as heavy smokers. Current smokers with missing data in the number of cigarettes smoked per day field (field #3456) were treated as moderate smokers. |
| Systolic blood pressure (SBP) | UKB:4080 & UKB:93 | |
| Standard deviation of SBP | UKB:4080 & UKB:93 | |
| Ratio of total cholesterol:HDL | UKB:30690 & UKB:30760 | |
| Family history of myocardial infarction before the age of 60 | NA | Set to FALSE for all participants, UK Biobank family history touchscreen questionnaire (field ID category #100034) did not include age of onset. |
| Diabetes status | PMID:27631769 | Defined using Eastwood et al. (2016) algorithm. |
| History of atrial fibrillation | UKB:20002 & HES | ICD-10 code I48, or ICD-9 codes 427.31 or 427.32 |
| Chronic kidney disease | UKB:42026 & HES | CKD stage 3 or higher (ICD-10 codes N18.3 or N18.4, or ICD-9 codes 585.3–585.6), nephrotic syndrome (ICD-10 code N04 or ICD-9 code 581), chronic glomerulonephritis (ICD-10 code N03 or ICD-9 code 582), chronic pyelonephritis (ICD-10 code N11 or ICD-9 code 590.0), dependence on renal dialysis (ICD-10 code Z99.2 or ICD-9 code V45.11), or history of kidney transplant (ICD-10 code Z94.0 or ICD-9 code V42.0). |
| History of severe mental illness | UKB:20002 & HES | ICD-10 codes F20, F31, or F32.1–F32.3, or ICD-9 codes 295, 296.0, 296.22–296.24, 296.4–296.7, or 298.0 |
| History of migraine | UKB:20002 & HES | ICD-10 codes G43 or G44.0, or ICD-9 codes 339.0 or 346 |
| Rheumatoid arthritis | UKB:20002 & HES | ICD-10 codes M05 or M06, or ICD-9 code 714 |
| Systemic lupus erythematosus | UKB:20002 & HES | ICD-10 code M32, or ICD-9 code 710.0 |

| Continuation of Table S6 | | |
|---|---|---|
| Risk factor | UKB field / source | Phenotyping details |
| Erectile dysfunction | UKB:20002 & UKB:2003 | Combination of self-report and related medications |
| Blood pressure lowering medication | UKB:6177 & UKB:6153 | As per list of medications listed in the QRISK3 publication |
| Systematic corticosteroid medication | UKB:20003 | As per list of medications listed in the QRISK3 publication |
| Atypical antipsychotic medication | UKB:20003 | As per list of medications listed in the QRISK3 publication |

Table S5: **List of medications used for sample exclusion criteria (statin-takers).**

| Medication code | Medication label |
|---|---|
| 1140861892 | acipimox |
| 1141146234 | atorvastatin |
| 1140861924 | bezafibrate |
| 1141157260 | bezafibrate product |
| 1140862026 | ciprofibrate |
| 1140888590 | colestipol |
| 1140909780 | colestyramine |
| 1141180734 | colestyramine product |
| 1141180722 | colestyramine+aspartame 4g/sachet powder |
| 1141192736 | ezetimibe |
| 1140861954 | fenofibrate |
| 1140888594 | fluvastatin |
| 1140861856 | gemfibrozil |
| 1141157262 | gemfibrozil product |
| 1140861868 | nicotinic acid product |
| 1140888648 | pravastatin |
| 1141192410 | rosuvastatin |
| 1140861958 | simvastatin |

Medication codes and labels are as given in UK Biobank field 20003.

Table S7: **Sex-rescaled QRISK3 scaling factors**

| Sex | Model | D1 | D2 | D3 | D4 | D5 | D6 | D7 | D8 | D9 | D10 |
|---|---|---|---|---|---|---|---|---|---|---|---|
| Male | QRISK3 | 0.529 | 0.715 | 0.727 | 0.743 | 0.715 | 0.771 | 0.748 | 0.713 | 0.782 | 0.726 |
| Female | QRISK3 | 0.602 | 0.786 | 0.710 | 0.684 | 0.641 | 0.734 | 0.630 | 0.676 | 0.699 | 0.712 |

Sex-based rescaling factors for QRISK3 presented for each risk decile (D1 to D10). The recalibrated model decile values are calculated by dividing the original risk value by the corresponding rescaling factor (rounded to 2 decimal places). These factors enable sex-calibration through sex-specific adjustment of predicted risks, facilitating fairer and more comparable risk predictions.

Table S8: **Calibration intercept and slope with 95% CI (3 decimals) for sex-recalibrated models**

| Strata | Fairness group | Recalibration | Calibration intercept | Calibration slope |
|---|---|---|---|---|
| Ethnicity | Asian or Asian British | Sex | 0.044 [-0.260, 0.349] | 0.946 [0.826, 1.066] |
| Ethnicity | Black or Black British | Sex | -0.371 [-0.926, 0.184] | 0.868 [0.693, 1.042] |
| Ethnicity | Other ethnic group | Sex | 0.159 [-0.216, 0.533] | 1.003 [0.875, 1.130] |
| Ethnicity | White | Sex | -0.059 [-0.110, -0.008] | 0.989 [0.969, 1.009] |
| IMD | Q1 | Sex | -0.035 [-0.152, 0.082] | 1.013 [0.969, 1.057] |
| IMD | Q2 | Sex | -0.117 [-0.232, -0.002] | 0.973 [0.929, 1.016] |
| IMD | Q3 | Sex | -0.064 [-0.177, 0.048] | 0.984 [0.941, 1.027] |
| IMD | Q4 | Sex | 0.026 [-0.084, 0.136] | 1.017 [0.975, 1.060] |
| IMD | Q5 | Sex | -0.137 [-0.239, -0.035] | 0.930 [0.889, 0.970] |

Table S9: **Fairness metrics for one-stage sex-recalibrated QRISK3 models with 95% confidence intervals.**

| Characteristic | AED | CUAED | EOD | TED |
|---|---|---|---|---|
| Ethnicity | 0.13 [0.12, 0.14] | 0.06 [0.02, 0.10] | 0.31 [0.23, 0.38] | 0.44 [0.30, 0.61] |
| Sex | 0.23 [0.23, 0.23] | 0.02 [0.01, 0.02] | 0.39 [0.38, 0.41] | 0.49 [0.47, 0.51] |
| Age | 0.35 [0.34, 0.35] | 0.04 [0.04, 0.04] | 0.57 [0.56, 0.58] | 3.46 [3.15, 3.83] |
| Imd | 0.06 [0.05, 0.06] | 0.02 [0.01, 0.03] | 0.08 [0.06, 0.11] | 0.04 [0.03, 0.06] |

Fairness metric values are presented for the sex-recalibrated QRISK3 model, for each characteristic of interest (ethnicity, sex, age, IMD, ethnicity&IMD, and ethnicity&sex). The metrics are Accuracy Equality Difference (AED), Conditional Use Accuracy Equality Difference (CUAED), Equal Opportunity Difference (EOD), and Treatment Equality Difference (TED), each reported as a mean with 95% confidence intervals (CI) across 200 bootstraps. Larger values indicate greater differences in model performance across groups.

Table S10: **Performance metrics for sex-recalibrated QRISK3 model alone, and differences in performance metrics between two-stage and one-stage model.**

| PRS Threshold | Accuracy | Precision | Sensitivity | Specificity |
|---|---|---|---|---|
| **QRISK3 (one-stage model)** | 0.817 [0.816, 0.818] | 0.140 [0.137, 0.143] | 0.434 [0.427, 0.441] | 0.84 [0.839, 0.841] |
| 1 | -0.002 [-0.002 , -0.002] | -0.00 [-0.000, -0.000] | 0.006 [0.005, 0.007] | -0.002 [-0.002, -0.002] |
| 2 | -0.004 [-0.004, -0.003] | -0.000 [-0.001, 0.000] | 0.011 [0.010, 0.013] | -0.004 [-0.005, -0.004] |
| 5 | -0.009 [-0.009, -0.009] | -0.001 [-0.002, -0.000] | 0.027 [0.025, 0.029] | -0.011 [-0.012, -0.011] |
| 10 | -0.019 [-0.020, -0.019] | -0.003 [-0.004, -0.002] | 0.050 [0.047, 0.052] | -0.023 [-0.024, -0.023] |
| 20 | -0.041 [-0.041, -0.040] | -0.009 [-0.010, -0.008] | 0.089 [0.085, 0.092] | -0.048 [-0.049, -0.048] |

Differences in accuracy, precision, sensitivity, and specificity between two-stage and one-stage versions of the recalibrated QRISK3 model, at each PRS percentile threshold (1%, 2%, 5%, 10%, and 20%). Differences are reported as mean values with 95% confidence intervals (CIs) derived from 200 bootstrapped samples. Negative values indicate a reduction in the performance metric for the two-stage recalibrated model relative to the one-stage recalibrated model; positive values indicate an increase.

Table S11: **Fairness metrics for one-stage sex-recalibrated QRISK3 model.**

| Strata | Metric | Bootstrap estimate | 95%CI lower | 95%CI upper |
|---|---|---|---|---|
| Age | AED | 0.347 | 0.344 | 0.351 |
| Age | EOD | 0.568 | 0.555 | 0.581 |
| Age | TED | 3.464 | 3.163 | 3.77 |
| Age | CUAED | 0.037 | 0.035 | 0.04 |
| Sex | AED | 0.232 | 0.229 | 0.235 |
| Sex | EOD | 0.393 | 0.38 | 0.405 |
| Sex | TED | 0.488 | 0.471 | 0.505 |
| Sex | CUAED | 0.016 | 0.014 | 0.02 |
| Ethnicity | AED | 0.132 | 0.119 | 0.145 |
| Ethnicity | EOD | 0.307 | 0.237 | 0.378 |
| Ethnicity | TED | 0.437 | 0.313 | 0.59 |
| Ethnicity | CUAED | 0.057 | 0.027 | 0.098 |
| IMD | AED | 0.057 | 0.053 | 0.061 |
| IMD | EOD | 0.082 | 0.063 | 0.102 |
| IMD | TED | 0.042 | 0.03 | 0.056 |
| IMD | CUAED | 0.018 | 0.01 | 0.025 |

Fairness metrics (AED, EOD, TED, CUAED) for baseline one-stage sex-recalibrated fairness metrics. Reported as mean values with 95% confidence intervals (CIs) derived from 200 bootstrapped samples.
